# Clinical and functional connectivity outcomes of 5-Hz repeated transcranial magnetic stimulation as an add-on treatment in cocaine use disorder: a double-blind randomized controlled trial

**DOI:** 10.1101/2020.07.15.20154708

**Authors:** Eduardo A. Garza-Villarreal, Ruth Alcala-Lozano, Sofia Fernandez-Lozano, Erik Morelos-Santana, Alan Dávalos, Viviana Villicaña, Sarael Alcauter, F. Xavier Castellanos, Jorge J. Gonzalez-Olvera

## Abstract

**BACKGROUND:** Cocaine use disorder (CUD) is a global condition lacking effective treatment. Repeated magnetic transcranial stimulation (rTMS) may reduce craving and frequency of cocaine use, but little is known about its efficacy and neural effects.

**OBJECTIVE/HYPOTHESIS:** We sought to elucidate short- and long-term clinical benefits of 5-Hz rTMS as an add-on to standard treatment in CUD patients and discern underlying functional connectivity (FC) effects using magnetic resonance imaging.

**METHODS:** Forty-four CUD patients were randomly assigned to complete the 2-week double-blind randomized controlled trial (acute phase) [Sham (n=20, 2 female) and Active (n=24, 4 female)], in which they received 2 daily sessions of rTMS on the left dorsolateral prefrontal cortex. Subsequently, 20 CUD patients continued to an open-label maintenance phase for 6 months (2 weekly sessions for up to 6 months).

**RESULTS:** rTMS plus standard treatment for 2 weeks significantly reduced craving (p = 0.013, d=0.77) and impulsivity (p = 0.011, d=0.79) in the Active group. We also found increased FC between lDLPFC-vmPFC and vmPFC-right angular gyrus Clinical and functional connectivity effects were maintained for 3 months but they dissipated by 6 months. We did not observe reduction of positive cocaine urine tests, however, self-reported frequency and grams consumed for 6 months were reduced.

**CONCLUSIONS:** With this RCT we show that 5-Hz rTMS has potential promise as an adjunctive treatment for CUD and merits further research.

## Introduction

Cocaine use disorder (CUD) is characterized by impulsivity and intense craving, with consequent risk-taking and antisocial behavior and health sequelae that can include violence, hospitalizations and death [1]. There are no approved medications for CUD and standard-of-care psychosocial treatments are associated with high dropout rates [2,3]. Accordingly, new treatment approaches are needed; neuromodulation is an attractive contender [4]. Repeated transcranial magnetic stimulation (rTMS) has produced positive clinical outcomes in substance use disorders (SUDs), including reduction of craving and use in CUD [5–12]. For example, 15 Hz rTMS to the left dorsolateral prefrontal cortex (lDLPFC) reduced cocaine use frequency [7]. Theta burst stimulation (TBS) studies have also found promising results [13–16]. However, only one randomized controlled trial (RCT) with double blind [9] and few using sham stimulation have been conducted [9,10,17]. Thus, conclusions regarding the efficacy of rTMS for CUD are premature yet encouraging.

The neurobiological substrates of the potential clinical benefits of rTMS also remain under investigation. Circuits involving lDLPFC and ventromedial prefrontal cortex (vmPFC) have been reported to be pathological in CUD [10]. Hypofunctioning frontostriatal functional connectivity (FC) in CUD has been supported by animal models [18], in which prelimbic cortex (putative DLPFC homolog) stimulation reduced cocaine-seeking behavior, and inhibition increased it [19]. Evidence suggests that both excitatory lDLPFC and inhibitory vmPFC stimulation could reduce CUD symptoms [20,21]. The utility of excitatory and inhibitory stimulation is consistent with the apparent opposing intrinsic FC of the lDLPFC and vmPFC, i.e., spontaneous fluctuations of the BOLD signal in lDLPFC and vmPFC are negatively correlated in healthy controls [22], which might reflect their opposing functional and cognitive mechanisms in CUD [23]. Studies in major depression have shown that lDLPFC and subgenual FC, which are also negatively correlated, predicts improvement to rTMS treatment [24,25], highlighting a possible parallel with lDLPFC-vmPFC FC in CUD.

Here, we sought to determine the clinical efficacy of adding excitatory lDLPFC rTMS in CUD to standard therapy and to potentially identify circuit-based clinical target engagement in a double-blind RCT with longitudinal magnetic resonance imaging.

## Methods and Materials

Full methodological details can be found in the Supplementary material. Briefly, we recruited cocaine users meeting inclusion and exclusion criteria (Table S1). Relevant inclusion criteria were: 1) age 18 - 50 years, and 2) high consumption cocaine use for at least 1 year. The study was conducted at the Clinical Research Division of the National Institute of Psychiatry in Mexico City, Mexico, and all procedures were approved by the Institutional Ethics Research Committee and registered (CEI/C/070/2016; ClinicalTrials.gov NCT02986438). All participants were fully informed and provided written informed consent, per the Declaration of Helsinki. Figure S1 shows the CONSORT flowchart with study enrollment, visits and attrition; Figure 1 shows study design in detail. Briefly, we performed a 2-week parallel group double-blind RCT (Sham/Active) named the *acute phase*, followed by an open-label trial of chronic maintenance treatment, the *maintenance phase* as an add-on to standard treatment (Table S2). Clinical and MRI data acquisition were obtained at Baseline (T0), 2 weeks (T1), 3 months and 6 months.

**Figure 1.**
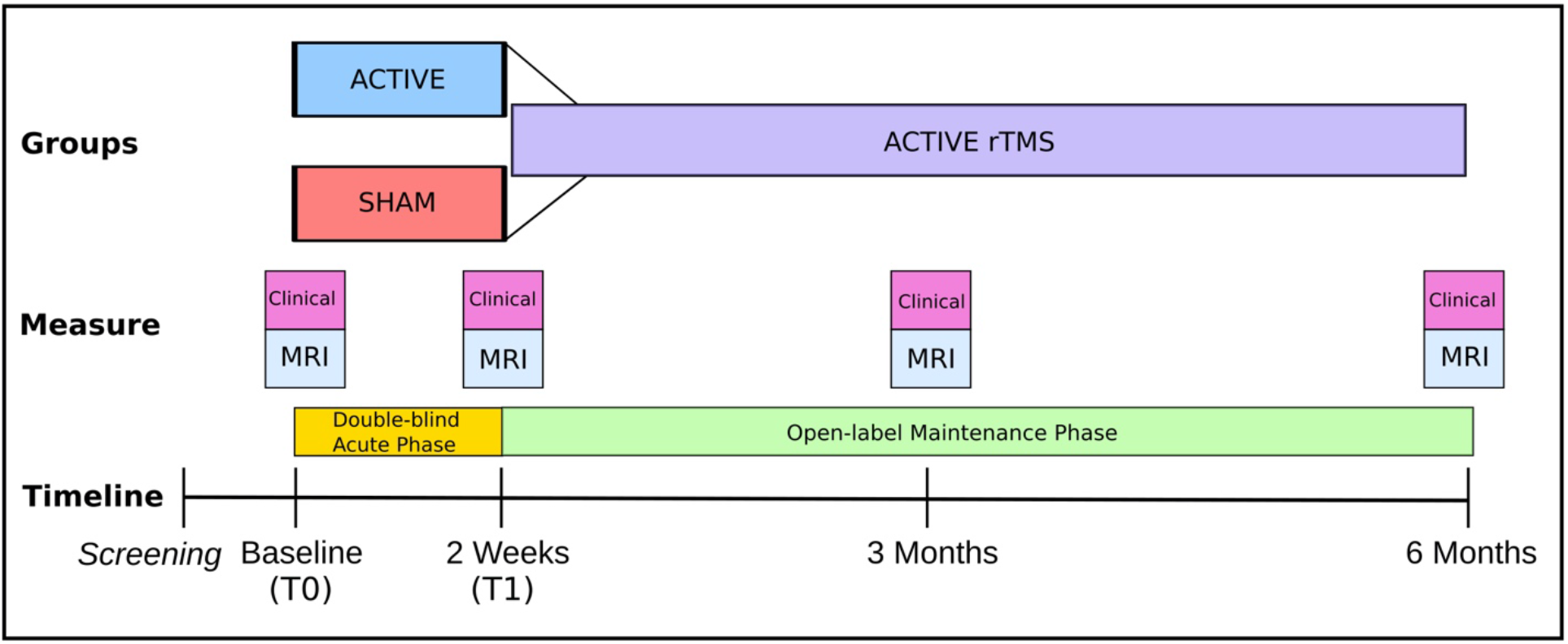
Design of the randomized clinical trial. MRI = Magnetic resonance imaging session; T0 = Time 0 or baseline; T1 = Time 1.

### Clinical assessments

For this study, primary outcome measures were 1) craving and 2) urine tests; secondary outcomes were: 1) impulsivity, 2) depression, 3) anxiety, and 4) sleep quality. Functional connectivity (FC) measured with resting state functional magnetic resonance imaging (rsfMRI) was a *tertiary* outcome to determine circuit-based clinical target engagement. Acute and maintenance phases were analyzed using linear Mixed Models and equivalent methods, to ascertain main and interaction effects.

### Transcranial magnetic stimulation

For the acute phase we used a MagPro R30+Option magnetic stimulator and a figure-eight B65-A/P coil (Magventure, Denmark); for the maintenance phase, we used a MagPro R30 stimulator and a figure-eight MCF-B70 coil (Magventure, Denmark) (reasons in Supplementary). During the 10 weekdays of the acute phase, 5,000 pulses per day were delivered at 5-Hz. The maintenance phase comprised two 5-Hz (5,000 pulses per day) sessions per week. Stimulation was delivered at 100% motor threshold to the lDLPFC, determined with standard clinical methods. Because a Brain Navigator was not available, we used a vitamin E capsule fiducial during MRI acquisition to identify the actual stimulation cortical target where rTMS was delivered (Fig. 2).

**Figure 2.**
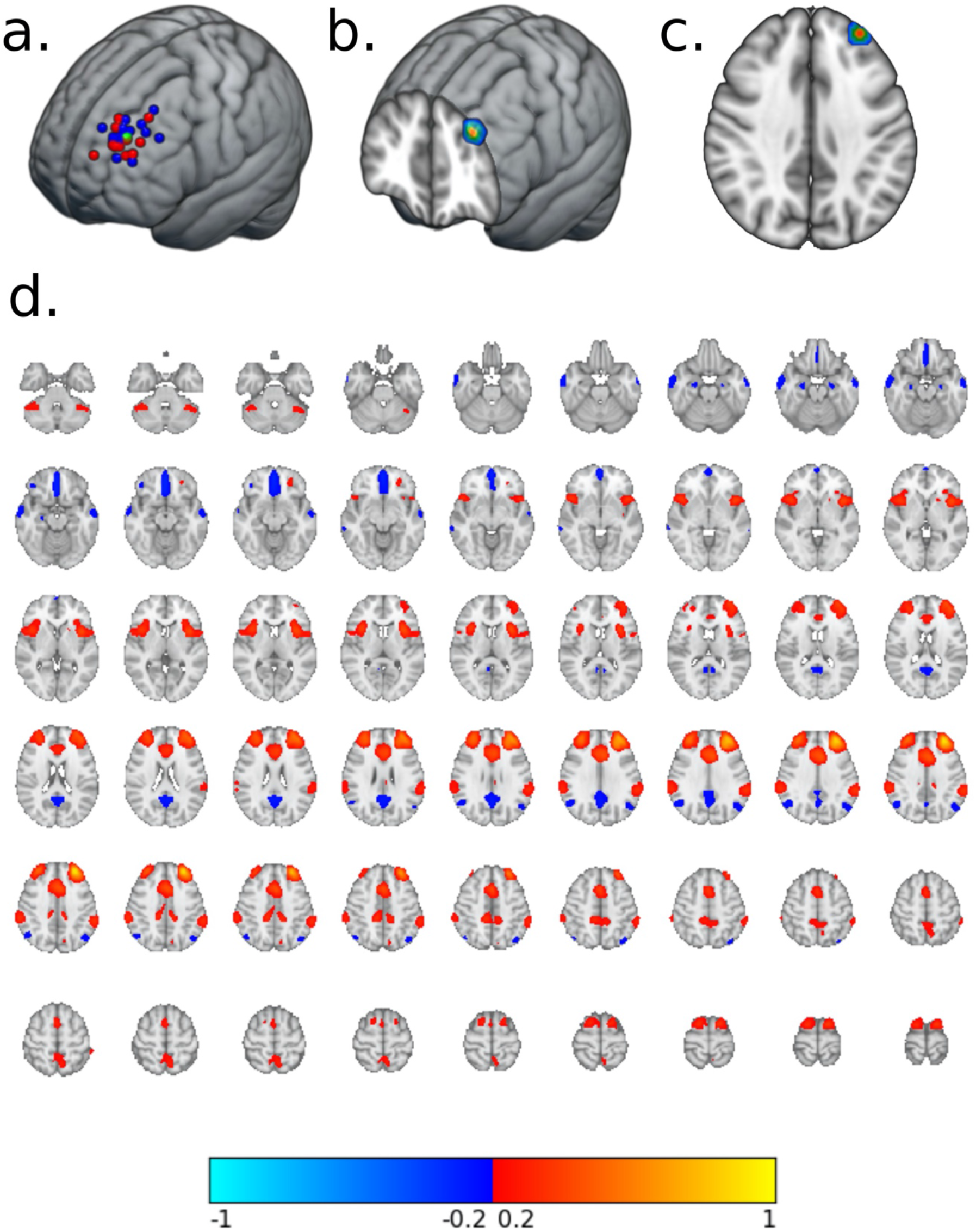
rTMS cortical targets and normative functional connectivity. a. Individual cortical rTMS targets of sham (red) and active (blue) groups, with the average target location (green); b. & c. Average target “cone” seed used to extract normative functional connectivity of the lDLPFC in the Human Connectome Project dataset (n = 1000); d. Normative functional connectivity map of the lDLPFC, from positive (red-yellow) to negative (blue-light blue) correlation.

### Magnetic resonance imaging

We acquired T1-weighted structural images and rsfMRI (10 min duration and TR=2s). We performed quality control and preprocessing using MRIQC version 0.15.2rc1 [26] and FMRIPREP version 1.5.5 [1, 2, RRID:SCR_016216]. Resting-state time series data from all participants and sessions were then processed using the XCP Engine v1.0 [27] with nuisance regression using the Power et al., pipeline [28]. Residual rsfMRI volumes were used for FC analysis. We first obtained normative lDLPFC FC following published methods [24] (Figure 2). The resulting normative lDLPFC FC map was clustered to obtain a vmPFC mask for ROI analysis (Table S4 & Fig. S3). We then calculated lDLPFC-whole brain FC in each subject and session, using the average lDLPFC cone mask. Because our primary interest was lDLPFC - vmPFC FC, the *acute phase* was analyzed with a 2 x 2 Mixed Model (group by session interaction) with the vmPFC mask as our ROI using FSL-randomise with Threshold-Free Cluster Enhancement and FWE correction. Afterwards, we explored the remaining clusters as ROIs with the same model. We then correlated the FC of significant clusters with significant clinical variables. The maintenance phase was analyzed extracting the mean FC from the resulting cluster or clusters in the acute phase analysis. Post-hoc, we calculated the individual FC of the significant vmPFC cluster from the acute phase analysis as the seed due to the importance of this region in SUDs and performed a whole-brain 2 x 2 mixed model analysis (group by session) of the acute phase and the maintenance phase.

## Results

The Sham and Active groups did not differ in demographic characteristics (Table 1), nor in baseline (T0) clinical measures (Table 2).

**Table 1.** Demographic characteristics between groups.

|  | Group |  | p |
| --- | --- | --- | --- |
|  | SHAM<br>(N=20) | ACTIVE<br>(N=24) |  |
| Sex |  |  | 0.84 |
| - Male | 18 (90.0%) | 20 (83.3%) |  |
| - Female | 2 (10.0%) | 4 (16.7%) |  |
| Age | 33.3 ± 8.7 | 35.8 ± 7.1 | 0.30 |
| Years of education | 12.9 ± 2.7 | 13.2 ± 3.0 | 0.79 |
| Marital status |  |  | 0.90 |
| - single | 13 (65.0%) | 13 (54.2%) |  |
| - married | 4 (20.0%) | 5 (20.8%) |  |
| - divorced | 1 ( 5.0%) | 3 (12.5%) |  |
| - separated | 1 ( 5.0%) | 2 ( 8.3%) |  |
| - widowed | 1 ( 5.0%) | 1 ( 4.2%) |  |
| Monthly income (MXN) | 5,080 ± 6,631 | 5,104 ± 6,910 | 0.99 |
| Employment (last 3 years) |  |  |  |
| - full time | 11 (55.0%) | 15 (62.5%) |  |
| - half time | 2 (10.0%) | 5 (20.8%) |  |
| - free lance | 5 (25.0%) | 4 (16.7%) |  |
| - student | 1 ( 5.0%) | 0 ( 0.0%) |  |
| - retired | 0 ( 0.0%) | 0 ( 0.0%) |  |
| - housewife | 0 ( 0.0%) | 0 ( 0.0%) |  |
| - unemployed | 1 ( 5.0%) | 0 ( 0.0%) |  |
| Main substance of use |  |  | 0.17 |
| - crack-cocaine | 17 (85.0%) | 24 (100.0%) |  |
| - cocaine | 3 (15.0%) | 0 ( 0.0%) |  |
| Onset age of cocaine use | 21.8 ± 6.8 | 23.4 ± 6.1 | 0.40 |
| Years of cocaine use | 10.1 ± 7.9 | 11.3 ± 7.9 | 0.61 |
| Socioeconomic status |  |  | 0.95 |
| - AB (High) | 2 (10.0%) | 2 ( 8.3%) |  |
| - C+ (Middle high) | 4 (20.0%) | 7 (29.2%) |  |
| - C (Middle) | 4 (20.0%) | 5 (20.8%) |  |
| - C- (Middle) | 5 (25.0%) | 4 (16.7%) |  |
| - D+ (Middle low) | 3 (15.0%) | 3 (12.5%) |  |
| - D (Low) | 2 (10.0%) | 2 ( 8.3%) |  |
| - E (Extreme poverty) | 0 ( 0.0%) | 1 ( 4.2%) |  |

*MXN = Mexican pesos, p = p-value, continuous normal variables = mean ± standard deviation, continuous non-normal variable = median and interquartile range, nominal = count (percentage from group). Statistical analyses are described in the methods section.*

**Table 2.** Baseline outcome measures per group.

|  | SHAM<br>(N=20) | Group<br>ACTIVE<br>(N=24) | p |
| --- | --- | --- | --- |
| Primary outcome measures |  |  |  |
| Craving: CCQ - General | 192.4 ± 47.9 | 187.2 ± 47.8 | 0.72 |
| Craving: CCQ - Now | 141.2 ± 49.4 | 148.5 ± 46.5 | 0.62 |
| Craving: VAS | 2.6 ± 2.8 | 3.9 ± 3.6 | 0.17 |
| Urine test: Cocaine |  |  | 0.89 |
| - positive | 7 (35.0%) | 10 (41.7%) |  |
| - negative | 13 (65.0%) | 14 (58.3%) |  |
| Secondary outcome measures |  |  |  |
| Impulsivity: BIS11 - Total | 60.8 ± 17.0 | 64.8 ± 16.8 | 0.44 |
| BIS 11 - Subdivisions |  |  |  |
| - Cognitive | 17.6 ± 5.4 | 18.2 ± 5.7 | 0.72 |
| - Motor | 18.9 ± 7.7 | 20.5 ± 7.8 | 0.49 |
| - Non-Planned | 24.2 ± 8.2 | 26.0 ± 6.0 | 0.42 |
| Anxiety: HARS score | 21.4 ± 15.8 | 17.8 ± 12.6 | 0.40 |
| HARS categories |  |  | 0.86 |
| - mild severity | 10 (50.0%) | 14 (58.3%) |  |
| - mild to moderate severity | 2 (10.0%) | 2 ( 8.3%) |  |
| - moderate-severe | 8 (40.0%) | 8 (33.3%) |  |
| Depression: HDRS score | 13.9 ± 10.2 | 13.8 ± 7.5 | 0.94 |
| HDRS categories |  |  | 0.85 |
| - normal | 8 (40.0%) | 7 (29.2%) |  |
| - minor depression | 2 (10.0%) | 2 ( 8.3%) |  |
| - less than major depressive | 3 (15.0%) | 7 (29.2%) |  |
| - major depression | 6 (30.0%) | 7 (29.2%) |  |
| - more than major depression | 1 ( 5.0%) | 1 ( 4.2%) |  |
| Sleep: PQSI total score | 10.7 ± 5.1 | 10.8 ± 4.2 | 0.95 |
| Global quality of sleep |  |  | 1.00 |
| - Good | 3 (15.0%) | 3 (12.5%) |  |
| - Bad | 17 (85.0%) | 21 (87.5%) |  |

*BIS 11 = Barrat Impulsiveness Scale, CCQ = Cocaine Craving Questionnaire, VAS = Visual analogue scale, HARS = Hamilton Anxiety Rating Scale, HDRS = Hamilton Depression Rating Scale, PQSI = Pittsburgh Sleep Quality Index, p = p-value.*

### Acute phase clinical assessment

Patients assigned to Active rTMS showed a significant reduction in craving visual analog scale (VAS) compared to Sham after 2 weeks of intensive treatment (group x time (GxT) interaction: t=-2.609, p=0.013, fdr=0.025, d=0.77) (Table 3). The effect on the Cocaine Craving Questionnaire Now section (CCQ-Now) did not reach significance (GxT interaction: t=-1.631, p=0.11, fdr=0.11, d=0.48), although VAS and CCQ-Now were significantly correlated at all time points (t=4.17, p=0.0002, r=0.54). Plots suggest that patients with high VAS at T0 had greater VAS reductions at T1 (Fig. 3). Impulsivity scores were also significantly reduced by Active compared to Sham stimulation (GxT: t=-2.677, p=0.011, fdr=0.042, d=0.79). Cocaine positive urine tests did not differ significantly; urine test results after 2 weeks of rTMS were: 1) Maintained negative (Sham n=12 [60%]; Active n=12 [50%]), 2) maintained positive (Sham n= 5 [25%]; Active=6 [25%]), 3) changed from negative to positive (Sham n=2 [10%]; Active n=3 [12.5%]), and 4) changed from positive to negative (Sham n=1 [5%]; Active n=3 [12.5%]) (see Table S8). Differently, anxiety, depression or sleep quality measures improved similarly and significantly in both groups after 2 weeks. Similar nonspecific results on quality of life (WHODAS) and broad psychopathology (SCL90-R) measures are shown in Figures S4 and Table S11.

**Table 3.** Outcome measures for the 2-week double-blind acute phase.

|  | Group |  |  |  |  |  |  |
| --- | --- | --- | --- | --- | --- | --- | --- |
|  | SHAM |  | ACTIVE |  |  |  |  |
|  | Time point |  |  |  | Group | Time point | Interaction |
|  | T0<br>(N=20) | T1<br>(N=20) | T0<br>(N=24) | T1<br>(N=24) |  |  |  |
| Primary outcome measures |  |  |  |  |  |  |  |
| Craving: CCQ - Now | 141.2 ± 49.4 | 129.2 ± 46.2 | 148.5 ± 46.5 | 115.2 ± 45.6 | t = 0.60, p = 0.55 | t = -1.26, p = 0.22 | t = -1.63, p = 0.11 |
| Craving: VAS | 2.6 ± 2.8 | 2.3 ± 2.5 | 3.9 ± 3.6 | 1.5 ± 2.4 | t = 1.63, p = 0.11 | t = -0.38, p = 0.71 | t = -2.61, p = 0.013* |
| Urine test: Cocaine |  |  |  |  |  |  | X<sup>2</sup> = 0.024, p = 0.88 |
| - positive | 7 (35.0%) | 9 (45.0%) | 10 (41.7%) | 10 (41.7%) |  |  |  |
| - negative | 13 (65.0%) | 11 (55.0%) | 14 (58.3%) | 14 (58.3%) |  |  |  |
| Secondary outcome measures |  |  |  |  |  |  |  |
| Impulsivity: BIS11 - Total | 60.8 ± 17.0 | 59.8 ± 21.4 | 64.8 ± 16.8 | 53.1 ± 17.4 | t = 0.79, p = 0.43 | t = -0.32, p = 0.75 | t = -2.68, p = 0.011* |
| BIS11 - Cognitive | 17.6 ± 5.4 | 18.2 ± 7.0 | 18.2 ± 5.7 | 14.4 ± 6.0 |  |  |  |
| BIS11 - Motor | 18.9 ± 7.7 | 19.4 ± 8.4 | 20.5 ± 7.8 | 15.8 ± 6.9 |  |  |  |
| BIS11 - Non-Planned | 24.2 ± 8.2 | 22.2 ± 10.3 | 26.0 ± 6.0 | 23.0 ± 6.4 |  |  |  |
| Anxiety: HARS score | 21.4 ± 15.8 | 11.7 ± 9.0 | 17.8 ± 12.6 | 10.0 ± 11.3 | t = -0.97, p = 0.34 | t = -3.15, p = 0.003* | t = 0.46, p = 0.65 |
| HARS categories |  |  |  |  |  |  |  |
| - mild severity | 10 (50.0%) | 15 (75.0%) | 14 (58.3%) | 21 (87.5%) |  |  |  |
| - mild to moderate severity | 2 (10.0%) | 2 (10.0%) | 2 ( 8.3%) | 1 ( 4.2%) |  |  |  |
| - moderate-severe | 8 (40.0%) | 3 (15.0%) | 8 (33.3%) | 2 ( 8.3%) |  |  |  |
| Depression: HDRS score | 13.9 ± 10.2 | 6.9 ± 7.5 | 13.8 ± 7.5 | 5.8 ± 5.5 | t = -0.23, p = 0.82 | t = -3.55, p = 0.001* | t = -0.32, p = 0.75 |
| HDRS categories |  |  |  |  |  |  |  |
| - normal | 8 (40.0%) | 15 (75.0%) | 7 (29.2%) | 16 (66.7%) |  |  |  |
| - minor depression | 2 (10.0%) | 2 (10.0%) | 2 ( 8.3%) | 6 (25.0%) |  |  |  |
| - less than major depressive | 3 (15.0%) | 1 ( 5.0%) | 7 (29.2%) | 0 ( 0.0%) |  |  |  |
| - major depression | 6 (30.0%) | 2 (10.0%) | 7 (29.2%) | 2 ( 8.3%) |  |  |  |
| - more than major depression | 1 ( 5.0%) | 0 ( 0.0%) | 1 ( 4.2%) | 0 ( 0.0%) |  |  |  |
| Sleep: PSQI total score | 10.7 ± 5.1 | 8.0 ± 2.9 | 10.8 ± 4.2 | 8.6 ± 3.4 | t = -0.14, p = 0.89 | t = -2.75, p = 0.009* | t = 0.43, p = 0.67 |
| Global quality of sleep |  |  |  |  |  |  |  |
| - Good | 3 (15.0%) | 5 (25.0%) | 3 (12.5%) | 4 (16.7%) |  |  |  |
| - Bad | 17 (85.0%) | 15 (75.0%) | 21 (87.5%) | 20 (83.3%) |  |  |  |

BIS 11 = Barratt Impulsiveness Scale, CCQ = Cocaine Craving Questionnaire, VAS = Visual analogue scale, HARS = Hamilton Anxiety Rating Scale, HDRS = Hamilton Depression Rating Scale, PSQI = Pittsburgh Sleep Quality Index, t = t-value, p = p-value, X<sup>2</sup> = Cochran-Mantel-Haenszel statistic, \* = statistically significant.

**Figure 3.**
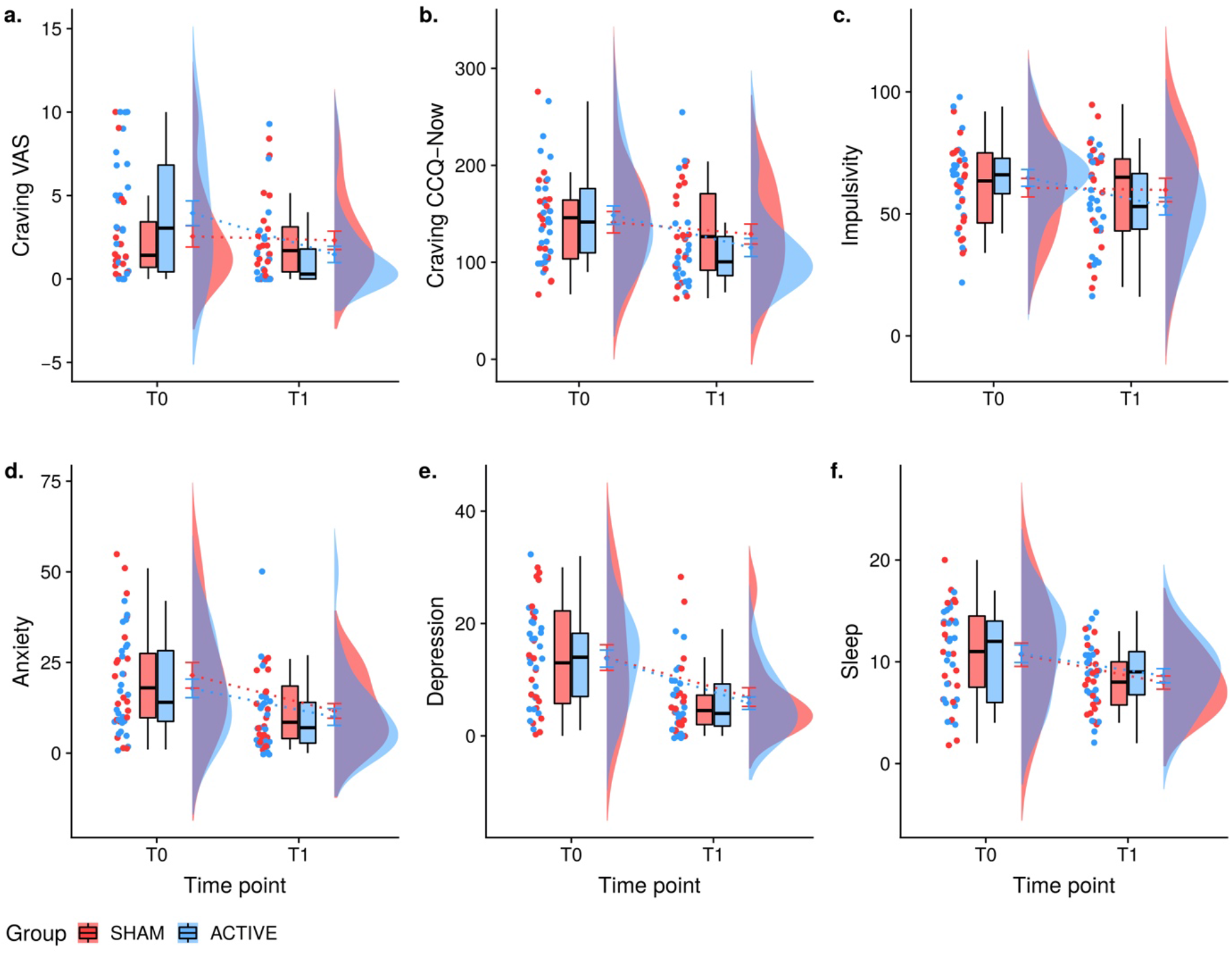
Raincloud interaction plots of the outcome measures for the 2-week double-blind acute phase. From left to right the plots shows at each time point: 1) the individual jittered points per measure per subject, 2) boxplots with median and whiskers without outliers, 3) means with error bars (SE) with dotted lines connecting each group’s mean between time points, and 4) flat violin plots showing the data distribution per group. T0 = Time 0 or baseline; T1 = Time 1 or 2 weeks; VAS = visual analog scale; CCQ = Cocaine Craving Questionnaire.

### Acute phase functional connectivity

We found a significant interaction in small vmPFC bilateral clusters (Table 4), in which the Active group showed higher lDLPFC-vmPFC FC than the Sham group at T1 (Fig. 4). The FC change did not correlate with any other measure. Post-hoc analysis showed a significant interaction between vmPFC-Right Angular gyrus (rAnG) FC, with higher FC in the Active group vs. Sham at T1 (Fig. 4). Our multiple regression showed that age (B = 0.02, t = 3.34, p = 0.004), sex (B = -0.15, t = -1.98, p = 0.064), years of consumption (B = -0.02, t = -3.31, p = 0.004) and CCQ-Now (B = -0.06, t = -1.91, p = 0.075) correlated significantly with the FC changes (adj-R^2^ = 0.28, p = 0.05). However, individually, CCQ-Now correlation was not significant (p = 0.075).

**Table 4.** Significant rsfMRI results in the acute double-blind phase.

| Seed region | Significant clusters | Cluster size | t-value | Peak coordinates (MNI) |  |  |
| --- | --- | --- | --- | --- | --- | --- |
|  |  |  |  | x | y | z |
| Left DLPFC | vmPFC (ROI) | 5 | 3.33 | -2.8 | 41.2 | -9.2 |
|  | vmPFC (ROI) | 2 | 4.41 | 6 | 41 | -10 |
| vmPFC | Angular gyrus (WB) | 265 | 3.28 | 56.3 | -45.2 | 42.3 |

ROI = region of interest, WB = whole brain.

**Table 4.**
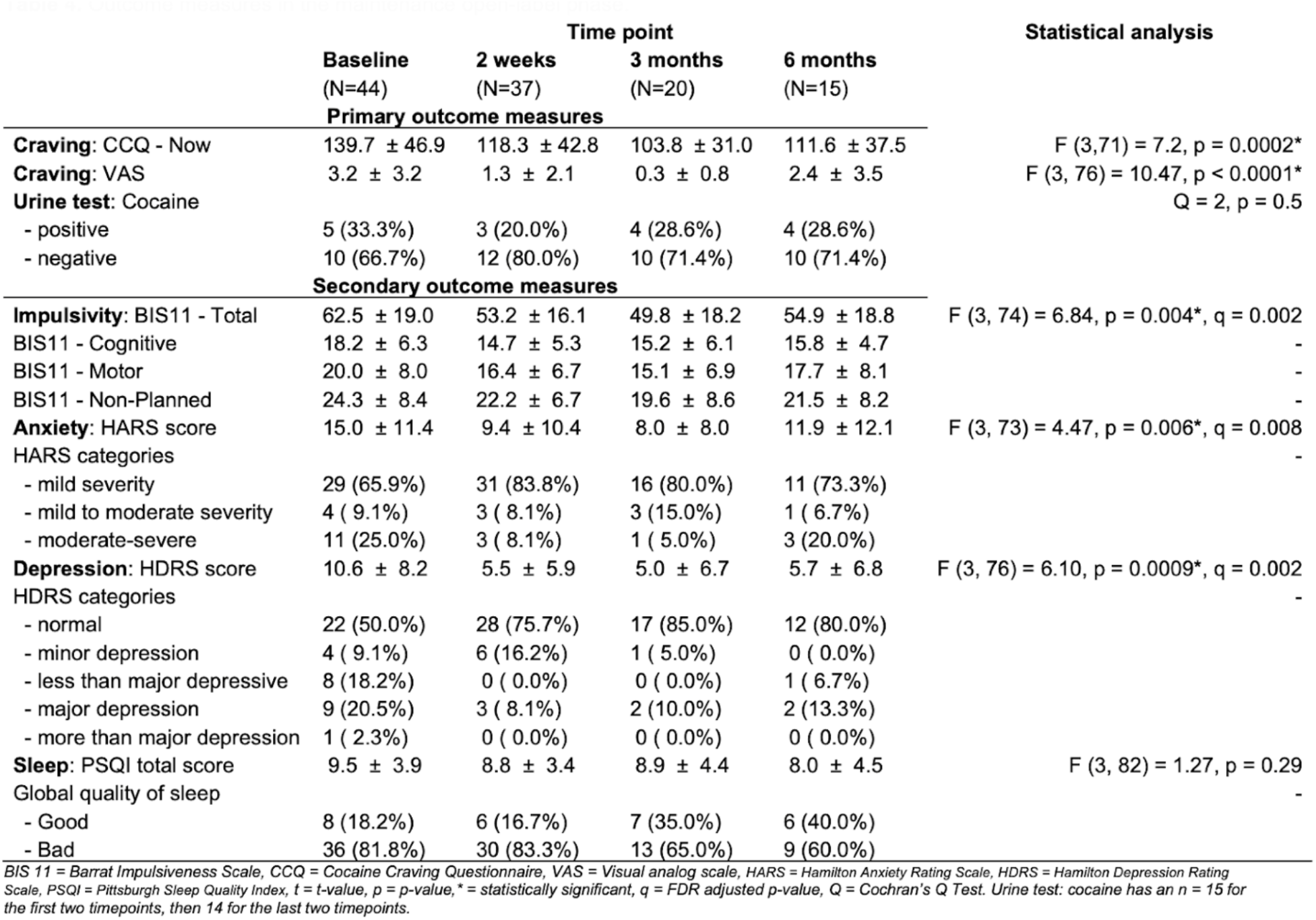
Outcome measures in the maintenance open-label phase.

**Figure 4.**
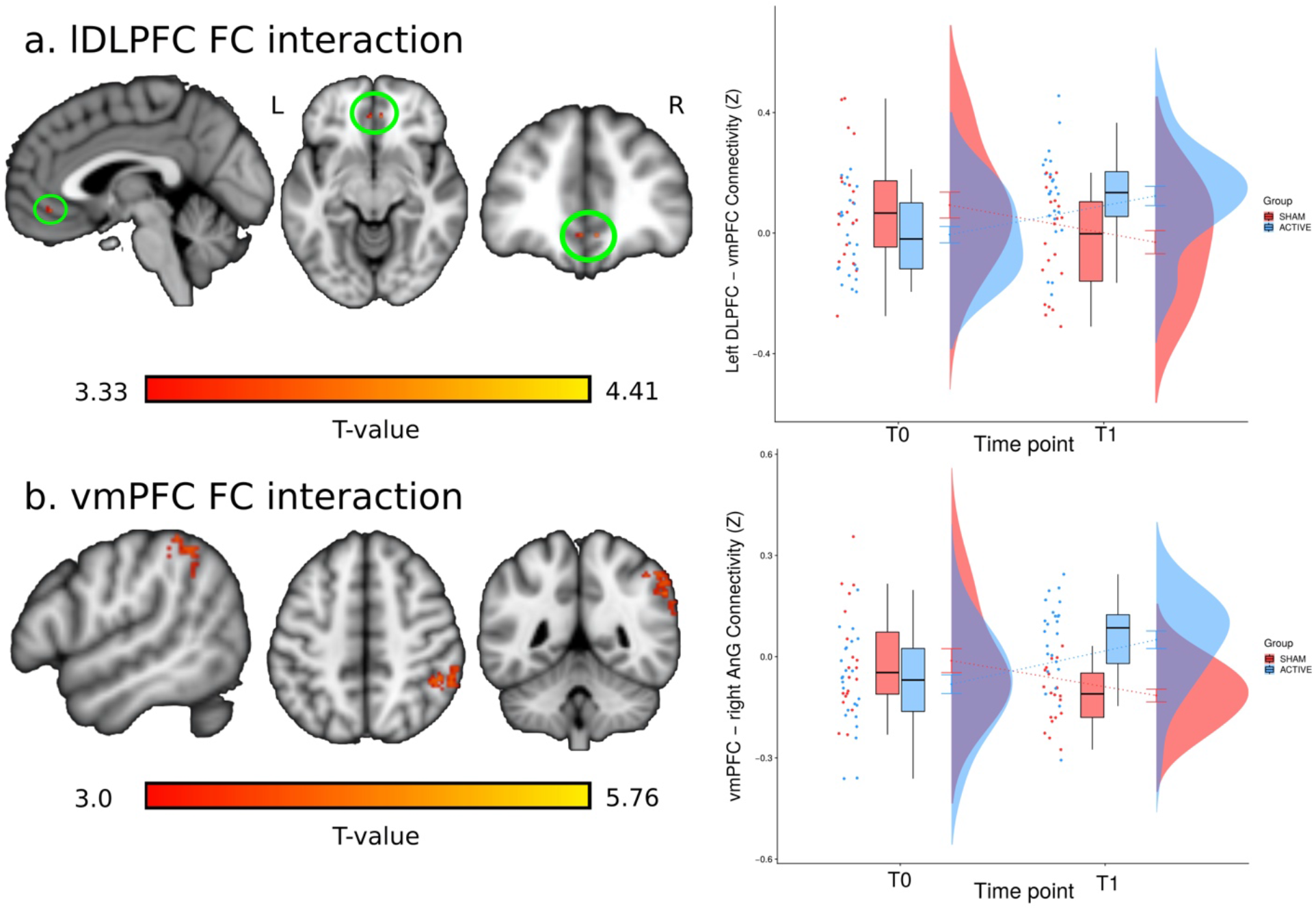
Functional connectivity results of the acute phase. a. lDLPFC-vmPFC significant interaction cluster; b. vmPFC-rAnG significant interaction cluster; right column shows the raincloud plots at each time point, where each data point represents the cluster’s mean Z-value for each subject; lDLPFC = left dorsolateral prefrontal cortex; vmPFC = ventromedial prefrontal cortex; rAnG = right angular gyrus; FC = functional connectivity.

### Maintenance phase clinical measures

We found significant reduction in both CCQ-Now and VAS craving measures after 6 months of open maintenance rTMS, but no significant effect on urine test results (Table 5 & Figure 6). Urine tests (n=14) were classified as: 1) Maintained negative (n=8 [53%]), 2) maintained positive (n=3 [10%]), 3) changed from negative to positive (n=1 [6.6%] at 6 months), and 4) changed from positive to negative (n=2 [13.3%] at 2 weeks) (see Tables S9). Post-hoc contrasts showed significant differences in CCQ-Now between BASELINE (T0) and all subsequent time points, but not among any other time points (Table S10). In craving VAS, contrasts showed differences between T0 and 2 WEEKS (T1)/3 MONTHS but not 6 MONTHS, with significant differences also noted between 3 MONTHS and 6 MONTHS. For the secondary outcome measures, we found significant differences in impulsivity, anxiety and depression, but not in sleep. Similarly to VAS, impulsivity, anxiety and depression differed between T0 and T1 and 3 MONTHS but not at 6 MONTHS. For details on statistical results, see Table S10. Figure S5 & Table S12 show similar results for WHODAS and SCL90-R. Finally, the modified Timeline Followback (mTLFB) indicated a significant reduction in frequency of use (X^2^ (6)=32.92, p<0.001, W=0.37) and grams of cocaine used (χ^2^(6)=36.57, p<0.001, W=0.41) (Fig. 7). *Post hoc* contrasts showed that this effect was driven only by higher frequency of use and grams consumed in the month before enrollment (Table S13 and S14).

**Figure 5.**
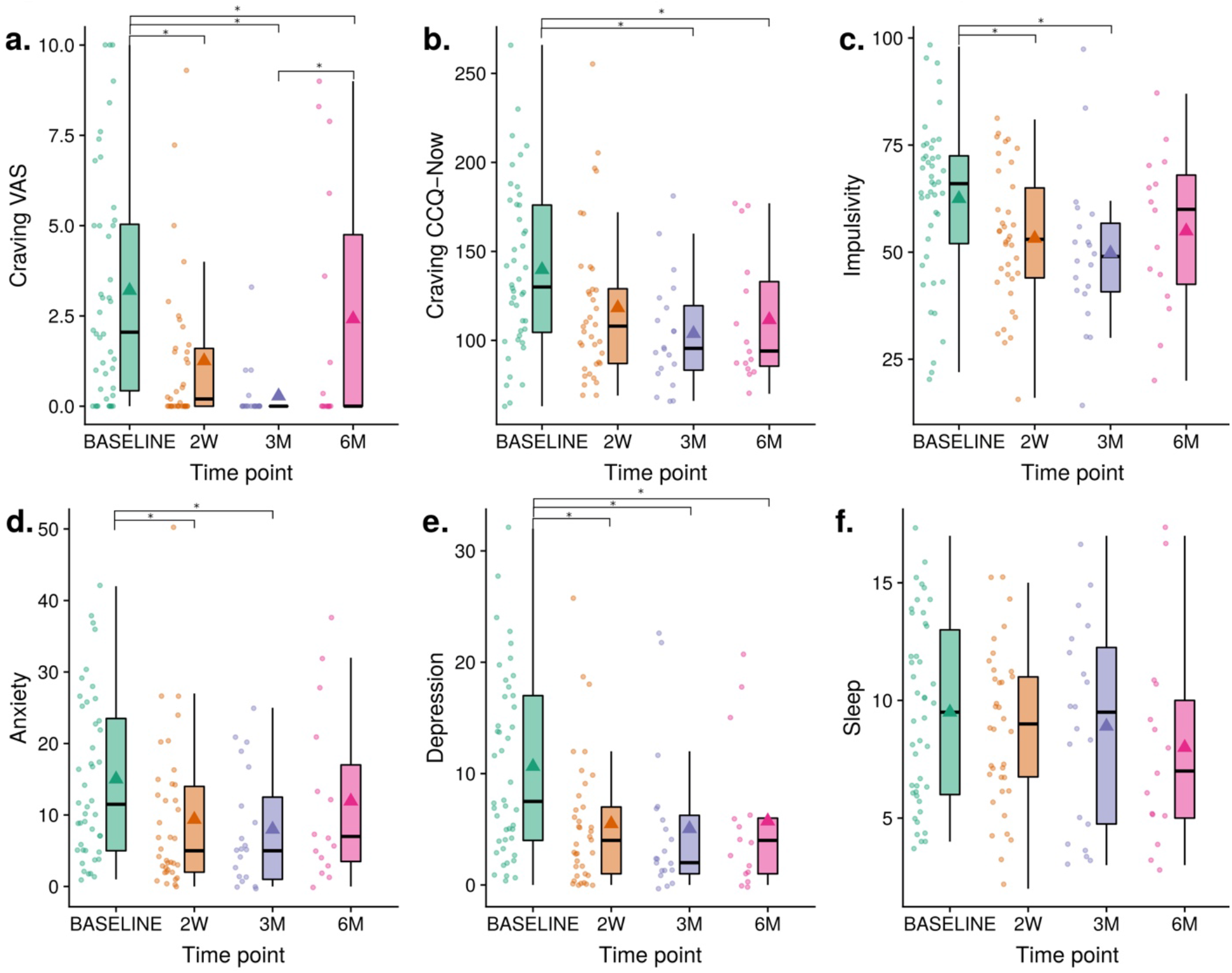
Plots of the outcome measures for the open-label maintenance phase. 2W = 2 weeks, 3M = 3 months, 6M = 6 months; Points to the left of each time point are the individual measures; boxplots show medians with whiskers and no outliers; ▲= mean; * = significant post-hoc comparison with Tukey correction.

**Figure 6.**
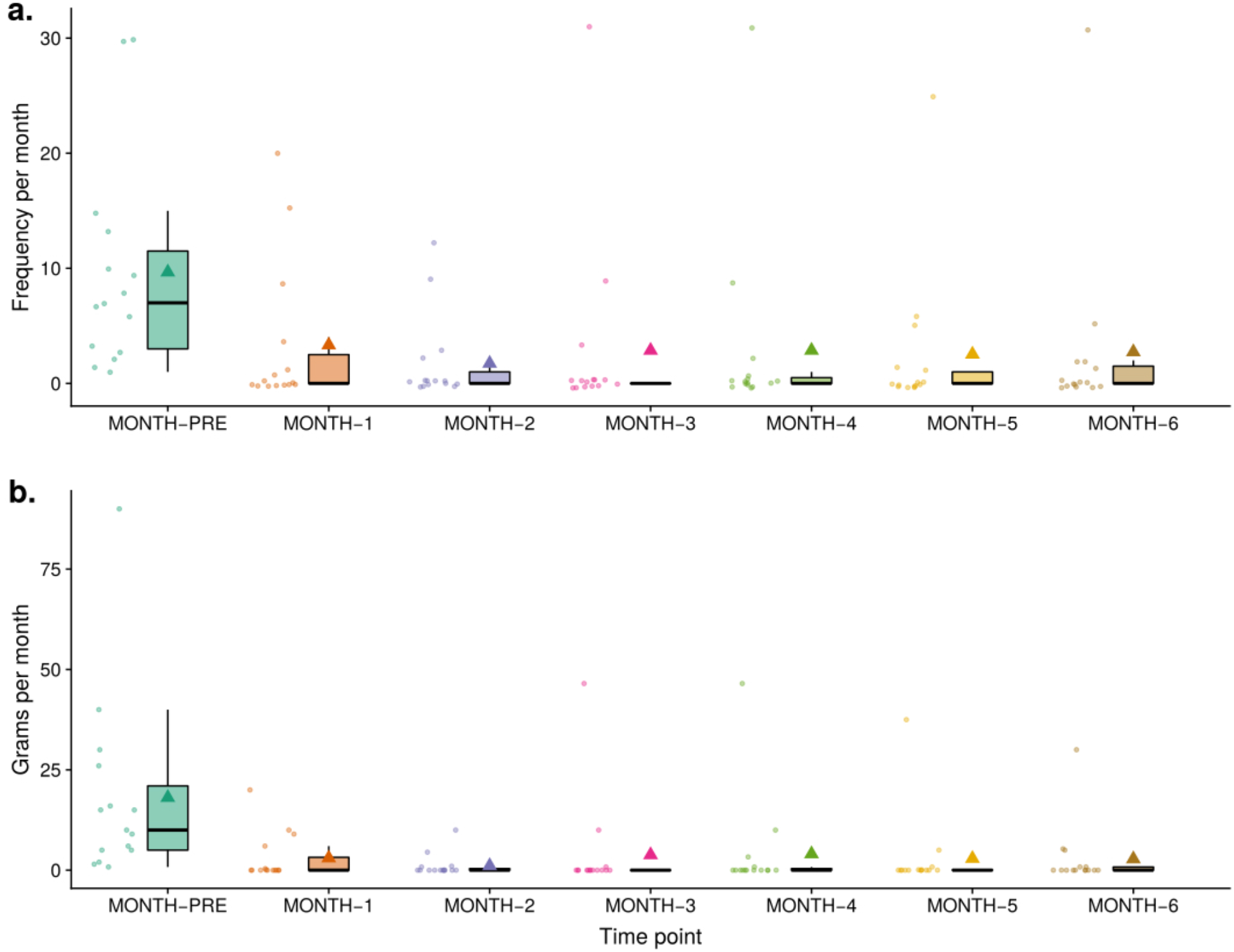
Modified Timeline Followback plots. MONTH-PRE = Previous month before the study started.

**Figure 7.**
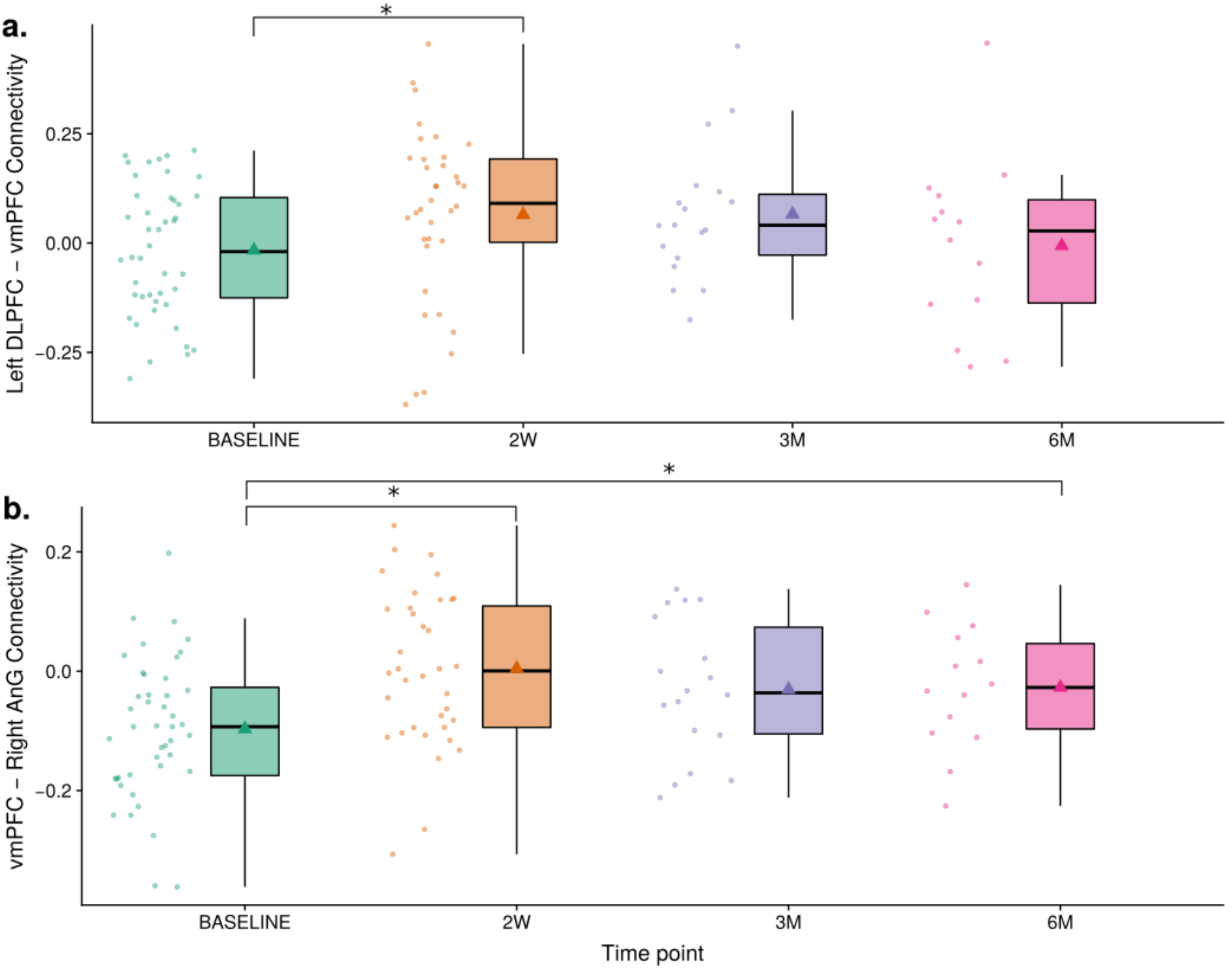
Functional connectivity in the open-label maintenance phase per cluster. 2W = 2 weeks, 3M = 3 months, 6M = 6 months; Points to the left of each time point are the individual measures; boxplots show medians with whiskers and no outliers;▲= mean; * = significant post-hoc comparison with Tukey correction; DLPFC = dorsolateral prefrontal cortex; vmPFC = ventromedial prefrontal cortex; AnG = angular gyrus.

### Maintenance phase functional connectivity

Mean FC of the lDLPFC-vmPFC cluster identified in the acute phase did not differ significantly over time (F(3, 79)=2.48, p=0.07); planned post-hoc analysis showed only the T0 vs. T1 contrast differed significantly (t=2.464, p=0.016). Mean FC of the vmPFC-rAnG cluster found in the acute phase differed significantly across time (F(3,72)=8.83, p<0.001); post-hoc analysis showed differences between T0 and T1 (t=5.12, p<0.001) and between T0 and 6 MONTHS (t=2.07, p=0.04) although the difference between T0 and 3 MONTHS did not reach significance (t=1.74, p=0.08) (Fig. 8).

## Discussion

Using a double-blind RCT, we found that 10 sessions of active rTMS over 2 weeks significantly reduced craving and impulsivity compared to sham, with the effect maintained during the open-label maintenance phase for 3 months. Effects did not remain statistically significant at 6 months, potentially because of decreased sample size/attrition. Other clinical symptoms, including anxiety and depression, improved similarly in both Active and Sham groups after 2 weeks, and were maintained for 3 months, returning to baseline at 6 months. Binary urine tests revealed no significant changes in cocaine use with treatment. Self-report on the modified Timeline Followback Method Assessment (mTLFB) showed lower frequency of cocaine use and grams used per month for 6 months. Functional connectivity (FC) analysis revealed that FC between left dorsolateral prefrontal cortex (lDLPFC) and ventromedial prefrontal cortex (vmPFC) increased in the Active rTMS group after 2 weeks, but the difference was not sustained during the lower intensity, open maintenance phase. lDLPFC-vmPFC FC did not correlate with impulsivity nor craving. Additionally, FC between vmPFC and right angular gyrus significantly increased after 2 weeks of active rTMS, with the effect largely sustained up to 6 months. This FC correlated sigificantly with age, years of cocaine consumption and sex, but not craving.

Overall, we found that: 1) 10 sessions of double-blind 5-Hz rTMS over two weeks had a greater effect on craving and impulsivity than placebo in CUD; 2) clinical effects on anxiety, depression and sleep quality were nonspecific to rTMS; and 3) the effectiveness of twice-a-week maintenance rTMS seemed to reduce by 6 months. A strength of our study is the double-blind RCT design for the first 2 weeks of intensive treatment, which revealed possible acute effects on craving and impulsivity of 5-Hz rTMS on the lDLPFC. These effects suggest that this scheme could be useful for CUD as an adjunct to standard treatments. Further research is warranted to determine how to best extend long term effects and to explore interactions with concomitant pharmacological and behavioral therapies. We now discuss our results and study limitations.

### Acute phase outcomes

Craving and impulsivity are subjective clinical symptoms that are commonly altered in CUD and are related to brain FC patterns [29,30]. Craving and impulsivity were reduced in our CUD patients after receiving Active rTMS compared to Sham. Previous studies on excitatory rTMS in CUD have found decreased craving [5,6,8,10]; similar results have been reported for alcohol, nicotine, methamphetamines and heroin [31–35]. Impulsivity is a multidimensional construct that is a marker of severity and a possible marker of treatment success in SUDs [36]. However, the only two prior studies on rTMS in SUDs that examined impulsivity did not find benefits when stimulating lDLPFC and mPFC respectively [37,38]. Hanlon et al. [21] suggested that lDLPFC stimulation in CUD would result in decreased impulsiveness in accordance with the competing neurobehavioral decision system (CNDS) model [23], by exciting the executive control network. The CNDS model also helps to explain reduced craving, whereby limbic system hyperactivity in CUD would be inhibited by increasing executive control network (ECN) activity, which is pathologically depressed in CUD. Overall, rTMS may reduce subjective craving and impulsivity by directly involving the ECN and the limbic system indirectly. By contrast, we did not find any differences in urine tests results, perhaps because of the low frequency of testing or the negative urine drug tests at baseline. Because all our patients were treatment-seeking, their initial commitment to the study may have allowed for the baseline negative urine tests.

Reduction or elimination of substance consumption is the ultimate goal for any SUD treatment. Nevertheless, reducing craving, impulsivity and other clinical comorbidities, as well as increasing quality of life, are also important in the treatment of addiction [39].

Contrary to our hypotheses, rTMS did not show greater benefits over standard treatment in anxiety, depression or sleep quality, as we found similar reduction in both Active and Sham groups after 2 weeks. Another study found improvement in sleep quality in CUD patients after rTMS therapy [40]. We also found improvements, but they were present in both groups, highlighting the importance of double-blind RCTs [40]. Although the lDLPFC is the main cortical target used in major depression, its clinical benefit is usually seen in patients resistant to pharmacological treatment after >3 weeks of 10 to 20-Hz rTMS therapy [41,42]. Moreover, studies in generalized anxiety disorder have shown clinical improvement in sham-controlled rTMS studies with right DLPFC inhibition (1-Hz) [43,44], but we again did not find benefit over standard treatment. The benefit of rTMS may be more present for longer periods of acute treatment.

We found that lDLPFC-vmPFC FC increased following 2 weeks of rTMS treatment only in the Active group, but the FC change was not significantly correlated with craving or impulsivity changes. Both regions are integral parts of the ECN, and previous studies have suggested the ECN is functionally depressed in CUD. These studies also suggest excitatory rTMS could reduce impulsivity by enhancing ECN function, while inhibitory rTMS to the vmPFC should decrease limbic circuit arousal, reducing craving [21,23].The relationship between these two regions seems to be important in CUD pathology and treatment response. An fMRI study using single-pulse TMS delivered to lDLPFC showed that stimulation decreased vmPFC activity in controls but not in cocaine users [10]. Speculatively, excitatory rTMS to the lDLPFC in CUD may functionally “correct” the lDLPFC-vmPFC circuit, however this would need to be investigated.

Excitatory rTMS over the lDLPFC has been found to increase low-frequency power of the BOLD signal in the vmPFC in healthy controls [45], and repeated stimulation may increase BOLD signal correlation between these two regions, such as our finding. The increased lDLPFC-vmPFC FC after 2 weeks of rTMS may be related to glutamatergic (Glut) activity. GABA dysregulation in the vmPFC appears to be related to cocaine seeking behavior [46]; increased GABA within vmPFC increases pathological limbic system activity by local inhibition [47,48]. Stimulating the altered lDLPFC circuit may result in local vmPFC increase of Glut activity (increased BOLD signal), counteracting GABA inhibition. However, this is speculative in the absence of spectroscopic examination and would need to be studied further.

Exploratoraly, we also found that vmPFC-rAnG FC increased in the Active rTMS group after 2 weeks, and this effect was also not correlated only with craving and impulsivity. The vmPFC and the AnG are both integral parts of the default-mode network (DMN) [49,50]. The vmPFC is part of the DMN medial temporal subsystem while the AnG is part of the DMN core, considered a cross-modal hub for information flow between subsystems, and involved in representing personally important information [51]. High AnG BOLD activation and concomitant low-levels of BOLD activation in the vmPFC during a Go-No Go response inhibition task predicted high levels of substance use and symptoms of dependency in adolescents [52]. Furthermore, the AnG has been correlated with craving scores in gaming and internet addiction [53–55]. The involvement of the AnG may also be in line with a preprint [56] that found that multi-day rTMS treatment increased FC across distinct non-targeted networks while it decreased FC within targeted networks [57]. rTMS stimulation of a given region has been hypothesized to induce compensatory network changes in global brain organization to maintain homeostasis [57]. Thus, our results suggest the DMN may be more involved in CUD than previously thought. [58].

### Maintenance phase outcomes

The positive clinical effect of rTMS seemed to decay over 6 months despite twice/week rTMS maintenance treatment in our study. We observed this in both primary and secondary outcome measures, except for sleep quality where there was no effect, as well as lDLPFC-vmPFC FC and other clinical measures (Supplementary Materials). By contrast, vmPFC-lAnG FC increased significantly at 2 weeks and was still elevated at 6 months. Our mTLFB assessment, which has been found to be reliable [59], showed significant reduction of monthly frequency of use and grams of cocaine used from the month prior to enrollment, though this monthly measure could only be obtained in the open-label maintenance phase due to the brevity of the acute phase. We lacked resources to study daily cocaine consumption, which remains an important goal for future work. This said, because there are no control subjects in this phase, it is impossible to disentangle the effect of rTMS and placebo effects.

In general, the clinical outcome of rTMS has been linked to three combined elements: 1) dosage, 2) frequency, and 3) specific region of stimulation. The number of rTMS pulses (dosage) may be related to the decay we observed, as they decreased from 25,000 pulses per week during the acute phase to 10,000 pulses per week in the maintenance phase. However, the number of pulses may not be related to clinical outcome, at least in depression [60] as there may be a ceiling effect for cortical excitability [61]. The range of frequencies with significant clinical improvement in CUD has not been directly explored and it will differ from region to region. In terms of the specific cortical target region, variation on the target within the lDLPFC is known to change clinical outcome in depression, due to its relationship with subgenual connectivity [24,25]. This has not been examined for SUDs. The DLPFC is a highly heterogeneous and functionally segmented area [62], for which both the 5.5 cm and the Beam F3 methods are prone to error [63,64]. Neuronavigation using individual FC is considered optimal for target specificity [65], however, it has not been proven to be clinically justified especially due to the cost of such a system and MRI, at least in developing countries. Another explanation for variability in treatment outcome may relate to cortical pathology, as cortical excitability of frontal regions in neuromodulation relies on the microstructural and neurochemical integrity of the cortical layers [66,67], which are altered in CUD [68–70]. Finally, the lack of long-term maintenance may reflect patient-level factors in CUD patients, such as their exposure to consumption triggers and other general treatment-related factors that need to be investigated further [2,3]. Treatment for SUDs is complex and any single strategy is unlikely to be sufficient to reduce consumption. Multiple therapeutic strategies may facilitate a reduction in cocaine consumption and other symptoms [3]. Nevertheless, here we show evidence for the possible benefit of rTMS to reduce craving, impulsivity and self-reported frequency and quantity of cocaine use, yet no effect on cocaine urine tests.

## Limitations

Full limitations are available in Supplementary materials. Briefly, study limitations include substantial dropout, which is common in the treatment of SUDs. A 12-month treatment study found an overall dropout rate of 31%, varying between 15 and 56% across subgroups [71]; the same group also found 30% dropout in CUD psychological treatment [72]. A 69% dropout rate for a 4-week treatment has also been reported in CUD [73]. Attrition in our study was: 1) 17% at 2 weeks; 2) 63% at 3 months; and 3) 72% at 6 months. The most common reasons were “poor adherence to treatment” followed by “incompatibility with working hours.” Overall, rTMS as an add-on treatment did not improve adherence beyond standard treatment. Motivation to remain in treatment should be measured in future studies to improve dropout rates. Moreover, all our CUD patients had to follow standard treatment, consisting of group psychotherapy, individual psychotherapy and pharmacotherapy. Several of our patients received pharmacotherapy concurrently with starting rTMS treatment but we did not find any differential effects of pharmacotherapies between active and sham groups. Our double-blind acute phase arm was limited to 2 weeks for ethical reasons, so as not to overextend placebo treatment for a vulnerable population. Nevertheless, that time window seemed sufficient to detect positive clinical effects. Future studies may benefit from using objective measures such as cue-reactivity [4]. Another factor may be our use of 5-Hz rTMS, which is in the low-end of excitatory frequencies, instead of 10-20 Hz, at the higher-end. However, 5- and 10-Hz produced similar clinical outcomes in major depression and Alzheimer’s disease [62,63]. We used 5-Hz to avoid secondary effects commonly seen in rTMS and increase treatment adherence. Finally, we cannot report on the effects of rTMS as a monotherapy for CUD. The overall goal of our study was to assess rTMS as an add-on therapy to the standard combination of CUD treatment as this is what clinicians encounter in their daily work. Despite these limitations, we were able to find a positive add-on effect of rTMS in the first 2 weeks and a possible maintenance effect, despite substantial attrition.

## Conclusions

In a double-blind RCT, we found that 5-Hz rTMS in cocaine addiction reduced subjective craving and impulsivity and increased functional connectivity between left dorsolateral prefrontal cortex with ventromedial prefrontal cortex (executive control network), and ventromedial prefrontal cortex with right angular gyrus (default mode network). The positive clinical and connectivity effects were higher in the acute, 2-week phase, and the maintenance effect (lower frequency of stimulation sessions) seemed to diminish after 3 months even with continued weekly therapy. However, craving and impulsivity did not correlate with functional connectivity changes. Standard treatment plus rTMS maintained a lower frequency of use and cocaine dosage than pre-baseline for 6 months during treatment, without concerns regarding the safety of rTMS. We conclude that rTMS therapy in cocaine addiction is a promising adjunctive treatment that may benefit treatment-seeking patients and that DLPFC-vmPFC and vmPFC-AnG are candidate circuit-based markers of clinical target engagement.

## Data Availability

Data and code are freely available. Main project website including code, data and statistical analyses: https://github.com/egarza/sudmex_tms_main; defaced raw data in BIDS format: https://openneuro.org/datasets/ds003037/versions/1.0.0; some derivatives: https://zenodo.org/record/3967527#.XzMDdXX0k5k; brain maps: https://neurovault.org/collections/8519/. More derivatives (i.e. fMRIprep outputs) available by request.

https://www.medrxiv.org/content/10.1101/2020.07.15.20154708v1

https://openneuro.org/datasets/ds003037/versions/1.0.0

https://zenodo.org/record/3967527#.XzMDdXX0k5k

https://neurovault.org/collections/8519/

## Open science initiative

Data and code are freely available. Main project website including code, data and statistical analyses: https://github.com/egarza/sudmex_tms_main; defaced raw data in BIDS format:

https://openneuro.org/datasets/ds003037/versions/1.0.0; some derivatives:

https://zenodo.org/record/3967527#.XzMDdXX0k5k; brain maps:

https://neurovault.org/collections/8519/. More derivatives (i.e. fMRIprep outputs) available by request.

## Acknowledgments

We would like to thank Alejandra Torres, Daniela Guerrero León, Ernesto Reyes Zamorano, Eden Sanchez Rosas, Hugo García Cantú and Isabel Espinoza Luna for their assistance in conducting the clinical trial. Major thanks to Michael D. Fox and Molly Schineller for providing the normative connectivity maps from fucidals. We also thank the Laboratorio Nacional de Visualización Científica Avanzada (LAVIS) for the use of their computer cluster and the Laboratorio Nacional de Imagenología por Resonancia Magnética (LANIREM). This study was supported by public funds CONACYT FOSISS No. 0260971 and CONACYT No. 253072. Student scholarships were provided by CONACYT for: Sofia Fernandez-Lozano No. 476284, Alan Dávalos No.581492 and Erik Morelos-Santana No. 479345.

