## Supplementary material for "Clinical and functional connectivity outcomes of 5-Hz repeated transcranial magnetic stimulation as an add-on treatment in cocaine use disorder: a double-blind randomized controlled trial"

### Supplementary Materials and Methods

**Study recruitment**

We recruited cocaine users according to inclusion and exclusion criteria (Table S1) via flyers to addiction and substance use disorder clinics and medical institutes in the Mexico City area, as well as through ads in social media. The study was conducted at the Clinical Research Division of the National Institute of Psychiatry in Mexico City, Mexico, and all procedures were approved by the Institutional Ethics Research Committee (CEI/C/070/2016). The trial was registered in ClinicalTrials.gov (NCT02986438). Before commencing any procedures, all participants were informed about the study and provided written informed consent, in line with the Declaration of Helsinki. Sample size was calculated using G*Power (1), for a 2 x 2 ANOVA with r = 0.3 (calculated from craving changes in previous cocaine rTMS studies), to attain 80% power at alpha = 0.05. All patients needed to be in psychosocial treatment for CUD, and most received medication. Types of treatments received during rTMS are in Table S2.

**Table S1.** Study criteria.

| **Inclusion** |
|---|

- Minimum age of 18 years and maximum of 50 years old.
- Cocaine use for at least 1 year, with current average use of at least 3 times a week, with periods of continuous abstinence of less than one month during the last year.
- Reading level of at least 6th grade of primary school.
- Ability to give valid informed consent.
- Right-handed (to avoid laterality bias).
- Body mass index ≦ 30.

| **Exclusion** |
|---|

- First-degree personal or family history of any clinically defined neurological disorder.
- Any electronic or metal implants or device (i.e., aneurysm clips, shunts, stimulators, cochlear implants, or electrodes, etc.).
- Splinters of metal or metal projectiles to the head or body.
- Current use of any investigational drug or of any medicine with anti- or pro-convulsive action such as tricyclic antidepressants or neuroleptics, unless prescribed for craving symptoms.
- History of schizophrenia, bipolar disorder, mania, or hypomania.
- History of any heart condition currently under medical care (i.e., myocardial infarction, angina pectoris, congestive heart failure, etc.)
- Women with reproductive potential not using an acceptable form of contraception, as well as pregnant or lactating women.
- Any history of seizures.
- Current dependence (by DSM-5 criteria) on substances other than cocaine and / or nicotine (cocaine use disorder).
- Claustrophobia.
- History of HIV infection or HIV antibody test positive (due to potential neuroinfection).

| **Elimination** |
|---|

- Expressed desire to stop participating.
- Those who for any reason stopped attending rTMS sessions, for 2 or more days for those in the acute phase, or 2 weeks for those in the maintenance phase.
- Those who presented abnormal radiological findings warranting clinical attention outside the study to ensure the health of the participant.
- The appearance of psychotic symptoms related to addictive disorder.
- Presence of adverse effects related to the application of rTMS such as seizures and abnormal elevation of mood.

*rTMS = repeated transcranial magnetic stimulation.*

**Table S2.** Standard treatments received by each participant during rTMS therapy.

| ID | Group | Received another treatment | Psychosocial treatment* | Pharmacological treatment | Number of medications | Medication 1 | Medication 2 | Medication 3 |
|---|---|---|---|---|---|---|---|---|
| 1 | Sham | yes | yes | yes | 2 | gabapentin | topiramate | NA |
| 2 | Sham | yes | no | yes | 2 | sertraline | clonazepam | NA |
| 5 | Sham | yes | no | yes | 1 | topiramate | NA | NA |
| 6 | Sham | no | no | no | 0 | NA | NA | NA |
| 8 | Sham | yes | yes | yes | 2 | fluoxetine | hydroxyzine | NA |
| 9 | Sham | yes | no | yes | 2 | citalopram | gabapentin | NA |
| 10 | Sham | yes | no | yes | 3 | fluoxetine | topiramate | hydroxyzine |
| 12 | Sham | yes | no | yes | 2 | atomoxetine | gabapentin | NA |
| 13 | Sham | yes | no | yes | 3 | escitalopram | topiramate | hydroxyzine |
| 18 | Sham | no | no | no | 0 | NA | NA | NA |
| 19 | Sham | yes | no | yes | 1 | oxcarbazepine | NA | NA |
| 22 | Sham | yes | no | yes | 2 | topiramate | paroxetine | NA |
| 27 | Sham | yes | yes | yes | 3 | sertraline | risperidone | valproic acid |
| 30 | Sham | yes | no | yes | 1 | topiramate | NA | NA |
| 33 | Sham | no | no | no | 0 | NA | NA | NA |
| 36 | Sham | yes | no | yes | 2 | topiramate | fluoxetine | NA |
| 40 | Sham | yes | no | yes | 2 | topiramate | citalopram | NA |
| 42 | Sham | yes | no | yes | 3 | topiramate | fluoxetine | atomoxetine |
| 45 | Sham | yes | no | yes | 3 | gabapentin | sertraline | hydroxyzine |
| 46 | Sham | yes | yes | yes | 3 | topiramate | venlafaxine | atomoxetine |
| 47 | Sham | no | no | no | 0 | NA | NA | NA |
| 50 | Sham | yes | no | yes | 3 | fluoxetine | topiramate | atomoxetine |
| 52 | Sham | yes | no | yes | 2 | fluoxetine | topiramate | NA |
| 3 | Treatment | yes | no | yes | 2 | topiramate | fluoxetine | NA |
| 4 | Treatment | yes | no | yes | 2 | bupropion | fluoxetine | NA |
| 7 | Treatment | no | no | no | 0 | NA | NA | NA |

| | | | | | | | | |
|---|---|---|---|---|---|---|---|---|
| 11 | Treatment | no | no | no | 0 | NA | NA | NA |
| 14 | Treatment | yes | no | yes | 1 | pregabalin | NA | NA |
| 15 | Treatment | no | no | no | 0 | NA | NA | NA |
| 16 | Treatment | yes | no | yes | 2 | mirtazapine | topiramate | NA |
| 17 | Treatment | yes | no | yes | 2 | topiramate | quetiapine | NA |
| 20 | Treatment | yes | no | yes | 1 | topiramate | NA | NA |
| 21 | Treatment | yes | yes | yes | 2 | topiramate | sertraline | NA |
| 23 | Treatment | yes | no | yes | 3 | gabapentin | hydroxyzine | mirtazapine |
| 24 | Treatment | yes | no | yes | 2 | fluoxetine | topiramate | NA |
| 25 | Treatment | yes | yes | yes | 4 | gabapentin | citalopram | atomoxetine |
| 26 | Treatment | yes | yes | yes | 2 | oxcarbazepine | quetiapine | NA |
| 28 | Treatment | yes | no | yes | 2 | fluoxetine | topiramate | NA |
| 29 | Treatment | yes | no | yes | 1 | topiramate | NA | NA |
| 31 | Treatment | no | no | no | 0 | NA | NA | NA |
| 32 | Treatment | no | no | no | 0 | NA | NA | NA |
| 34 | Treatment | yes | no | yes | 2 | topiramate | NA | NA |
| 35 | Treatment | yes | no | yes | 3 | topiramate | fluoxetine | atomoxetine |
| 37 | Treatment | yes | no | yes | 1 | topiramate | NA | NA |
| 38 | Treatment | yes | no | yes | 3 | topiramate | mirtazapine | NA |
| 39 | Treatment | yes | no | yes | 2 | topiramate | trazodone | NA |
| 41 | Treatment | yes | no | yes | 3 | topiramate | citalopram | gabapentin |
| 43 | Treatment | yes | no | yes | 2 | fluoxetine | topiramate | NA |
| 48 | Treatment | yes | yes | yes | 3 | fluoxetine | topiramate | hydroxyzine |
| 49 | Treatment | yes | no | yes | 2 | fluoxetine | topiramate | NA |
| 51 | Treatment | yes | no | yes | 3 | topiramate | fluoxetine | NA |
| 53 | Treatment | yes | yes | yes | 2 | valproic acid | quetiapine | NA |
| 54 | Treatment | yes | yes | yes | 3 | fluoxetine | topiramate | atomoxetine |

*The psychosocial treatment consisted of group therapy with a motivational approach focused on addiction, received at the addiction clinic of the National Institute of Psychiatry.*

**Study attrition**

Of the 54 recruited patients, 30 were randomly allocated to active treatment and 24 to sham rTMS (Figure S1). Five patients assigned to active rTMS and four assigned to sham discontinued the study, leaving 25 patients in the Active group and 20 in the Sham group who completed the acute phase. Following the double-blind phase, 14 patients in the Sham group opted for compassionate use and received 2 weeks of acute phase rTMS therapy. In the maintenance phase: 1) 20 patients (15 initially allocated to Active and 5 to Sham) finished 3 months of twice-weekly rTMS sessions (T2); 2) 15 patients (initially 10 Active and 5 Sham) finished 6 months of rTMS sessions (T3); and 3) 7 patients (initially 3 Active and 4 Sham) finished 12 months of twice-weekly rTMS sessions (T4). None of the patients who discontinued the study at any point reported adverse effects from rTMS as their reason. Due to substantial attrition at T1 (2 weeks), when the study was at ~30% completion we changed the maintenance phase to last 3 months instead of 12 months for new participants after approval by the ethics committee. Data collected up to the 6-months visit were analyzed due to the small sample size at 12 months (n = 7).

**Figure S1.** CONSORT flow diagram.

CONSORT
TRANSPARENT REPORTING of TRIALS

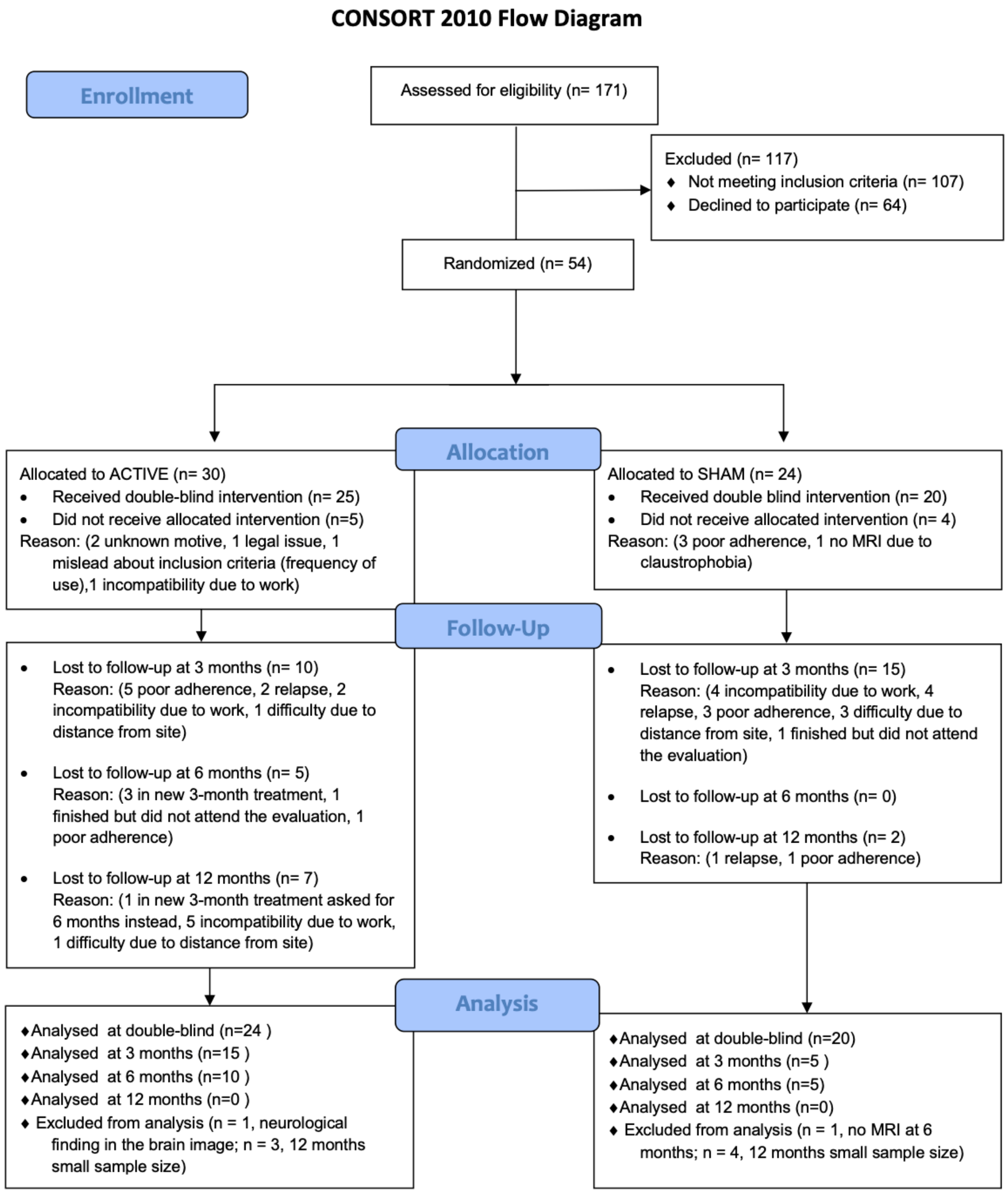

**Study timeline**

Patients were phone screened by RAL and AD (Figure S1). At Visit 1, screened patients arrived for a clinical screening interview by RAL and AD to confirm they met criteria. At Visit 2, enrolled patients underwent a full clinical assessment (Time 0 or T0). Initial MRI scanning occurred at Visit 3 (Baseline or MRI-T0). The clinical interview preceded MRI acquisition and always occurred within 3 days. Following MRI acquisition, we initiated the double-blind rTMS/sham *acute phase* (see below). Patients underwent regularly scheduled sessions (Active or Sham rTMS) for 10 days over 2 weeks. At the conclusion of 2 weeks (Visit 4; T1), they underwent clinical assessment and repeated MRI scanning, marking the end of the acute phase and the start of the open-label *maintenance phase*. The blind (active vs. sham) was decoded for each participant at the end of their acute phase. Patients assigned to Active rTMS entered the maintenance phase directly after T1. Patients assigned to Sham rTMS were given the choice to leave the study or continue with active open-label rTMS for compassionate use. Patients assigned to the Sham group who agreed to continue, received 2-weeks (10 days) acute treatment before continuing to the maintenance phase. The maintenance phase was initially designed to include 2 weekly rTMS sessions and clinical assessments and MRI scans at 3 months, 6 months and 12 months. However, the maintenance phase was subsequently changed to 3 months for new enrollments (see study attrition).

**Clinical Assessments details**

The following instruments were used in the overall clinical trial:

1. **MINI-PLUS**: Is a structured diagnostic interview, of short duration in which the main psychiatric disorders of Axis I of DSM-V and ICD-10 are explored for detection and / or diagnostic orientation, It is divided into modules, identified by letters, each corresponding to a diagnostic category. At the beginning of each module (except in the psychotic disorders module), the interview has one or more "filter" questions corresponding to the main diagnostic criteria for the disorder. At the end of each module, one or more diagnostic boxes are presented that allow the interviewer to indicate whether or not the diagnostic criteria for the disorder were met. This instrument will be used for the initial evaluation of the patient and verification of the inclusion and exclusion criteria (2).
2. **SCID-II**: Evaluate personality disorders in a categorical way according to DSM-IV criteria. Each of the criteria is valued from the following score: 1: absent, 2: Present or true, it

consists of 119 questions with a dichotomous answer that reduces the test administration time, The test was applied only in the baseline measurement (T0), since it is a constant clinical feature (3).

3. **SCL90 R**: The SCL-90-R is a self-applied symptom questionnaire consisting of 90 items. Each item is answered on a 5-point Likert-type scale, from "0" (absence of the symptom) to "4" (total presence of the same). By correcting the test we obtain 9 symptomatic scales and 3 indexes of psychological distress. The symptomatic scales are as follows: Somatization, Obsession-compulsion, Interpersonal sensitivity, Depression, Anxiety, Hostility, Phobic anxiety, Paranoid ideation and Psychoticism. The discomfort indices are: a) the global severity index (GSI), b) the positive symptomatic discomfort index (PSDI) and c) the total of positive symptoms (PST). The test was applied in each clinical measurement (T0 to T4) to assess changes in symptoms in each phase (4).
4. **Addiction Severity Index (ASI)**: The ASI is a semi-structured interview designed to address seven potential problem areas in substance-abusing patients: medical status, employment and support, drug use, alcohol use, legal status, family/social status, and psychiatric status. In 1 hour, a skilled interviewer can gather information on recent (past 30 days) and lifetime problems in all of the problem areas. The ASI provides an overview of problems related to substance, rather than focusing on any single area. The test was applied in each clinical measurement (T0 to T4) to assess changes in symptoms in each phase (5).
5. **BIS11**: The 11th version of the Barratt Impulsivity Scale is one of the most widely used instruments for assessing impulsivity. Its application is self-administered and it consists of 30 questions, grouped into three subscales: Cognitive impulsivity, Motor impulsiveness, Unplanned impulsiveness. Each of the questions has 4 possible answers (rarely or never, occasionally, often and always or almost always. The total score is the sum of all the items and the total of the subscales are the sum of the items corresponding to each of them (6).
6. **Hamilton Depression Rating Scale (HDRS)**: The Hamilton Rating Scale for Depression was used to provide a measure of the severity of depression. The version we used is the one of 17 items, recommended by the United States National Institute of Mental Health. Its content focuses on the basic aspects and behavior of depression, with vegetative, cognitive and anxiety symptoms having the greatest weight in the total calculation of the scale. The cutoff points to define severity are: no depression (0-7); mild depression (8-16); moderate depression (17-23); and severe depression (≥24). This scale was applied

in the basal measurement (T0) and all subsequent measurements. The test was applied in each clinical measurement (T0 to T4) to assess changes in symptoms in each phase (7).

7. **Hamilton Anxiety Rating Scale (HARS)**: This scale assesses the severity of anxiety globally and is useful for monitoring response to treatment. It is made up of 14 items, with 13 references to anxious signs and symptoms and the last one that evaluates the patient's behavior during the interview. The interviewer scores from 0 to 4 points each item, assessing both its intensity and frequency. The total score is the sum of those of each of the articles. The range is from 0 to 56 points. The optimal HAM-A score ranges were: no/minimal anxiety ≤ 7; mild anxiety = 8-14; moderate = 15-23; severe ≥ 24. The test was applied in each clinical measurement (T0 to T4) to assess changes in symptoms in each phase (8).
8. **Pittsburgh Sleep Quality Index (PSQI)**: This instrument has been created to measure the quality of sleep in patients with psychiatric disorders. It is made up of 24 items, although only 19 are taken into account for its correction. In addition, it is divided into 7 dimensions: Subjective sleep quality, Sleep latency. Duration of sleep, Usual sleep efficiency, Sleep disturbances, Use of medication, Daytime dysfunction. It is answered with a Likert-type scale that goes from 0 to 4. For its correction, a sleep profile is obtained in each of the dimensions ranging from 0 to 3 and a total score that can range from 0 to 21. The test was applied in each clinical measurement (T0 to T4) to assess changes in symptoms in each phase (9).
9. **Treatment-As-Usual follow-up**: Consists of a record of the treatment that each subject had indicated at the beginning of the study, which was prescribed by the treating physician in the addiction clinic of the National Institute of Psychiatry, according to the protocols that they normally follow. The record indicated whether the subject received psychotherapy and/or pharmacological treatment, together with the type of psychotherapy and the name of the drug received, as well as changes to these treatments in each of the following measurements. This record was made in a format created for the present study which was applied both in the baseline assessment and in each of the subsequent assessments.
10. **Timeline Followback Method Assessment modified (mTLFB)**: This is a record of the pattern of cocaine/crack use of each subject, made on a calendar-based format, where the consumption of the last two years up to the present was evaluated, indicating the number of days of use per month and the amount in grams consumed each full month (30 days). This format was applied in the baseline measurement (T0) where previous

consumption was recorded and in each subsequent measurement to assess the longitudinal pattern of substance use every month before and during the trial (10).

11. **Cocaine Craving Questionnaire (CCQ)**: This instrument evaluates the intensity of cocaine craving. The version used in this study has a format that evaluates craving at the present time, and a format that evaluates the general state of craving during the last week. Each form consists of 45 items, each item is made up of a 7-point Likert scale in which the subject must indicate their degree of agreement or disagreement, with some items scored inversely. For its interpretation, the total of the items is added. The test was applied at each clinical measurement (T0 to T4) to assess changes in craving in each phase (11).
12. **Cocaine Craving visual analogue scale (VAS)**: This is an instrument for the subjective evaluation of the subject's craving at the present moment. The visual scale consists of a continuous 100 mm line, in which the left endpoint refers to "no craving" and the right endpoint "the most intense craving" and the subject must mark with a cross the intensity of their craving at that moment between one of the two extremes. This scale was applied in each clinical measurement (T0 to T4) to assess changes in craving in each phase (12).
13. **Alcohol breath test**: An alcohol monitoring test was performed to identify the possible presence of substances in the subject before performing the MRI study. This was done in the initial evaluation (T0) and in each subsequent measurement (T1 to T4), with a breath alcohol analyzer, Lifeloc model FC10 (Wheat Ridge, CO, USA), which has a detection accuracy of ± .005 BAC.
14. **Urine drug test**: Performed to identify the possible presence of substances of abuse in subjects prior to performing the MRI study. This test was performed with a Kabla (Monterrey, NL, Mexico) reagent strip device, model Instant view-Drug screen, using the lateral flow chromatographic immunoassay technique. The substances detected and their cut-off points are as follows: Amphetamines (1000 ng/mL), Benzodiazepines (300 ng/mL), Cocaine (300 ng/mL), Methamphetamine (1000 ng/mL), Morphine/Opiates (2000 ng/mL), Marijuana/Hashish (50 ng/mL). This test was applied in the baseline measurement and each of the subsequent ones. Results in Tables S6 & S7.
15. **Reincidence/Relapse follow-up**: A record of the cocaine abuse patterns of patients was carried out, to identify if they presented reincidence or relapses. This was applied in each of the subsequent measurements (T1 to T4). "Reincidence" was defined as the presence of at least one episode of consumption but without returning to previous consumption, and "relapse" was defined when consumption returned to the previous pattern.

16. **WHODAS**: Instrument that assesses the psychological and social functioning of people affected by a mental disorder. It provides information on four areas: Personal Care, Occupation, Family/Housing and Social Functioning. The clinician scores to what extent there is a degree of deterioration in the interviewed person through a visual analog scale, which goes from 0 (absence of deterioration) to 5 (great deterioration). It is a descriptive scale that provides a total score and scores in each of the 4 dimensions. There are no cut points; the higher the score, the greater the disability. It was obtained in the baseline evaluation (T0) and in each of the subsequent ones (T1 to T4) (13).
17. **Edinburgh Handedness**: The Edinburgh Manual Laterality Inventory aims to assess manual dominance. This instrument evaluates the degree to which the subject uses the left or right hand for 4 predetermined actions and provides a numerical result, which is used to form three categories: predominant use of the left hand, similar use of both hands, and predominant use of the right hand. This instrument was applied in the baseline assessment (T0) only (14).

**Clinical outcome measures**

- Primary Outcome Measures:

1. Change in Cocaine Craving (CCQ) [ Time Frame: Baseline, 2 weeks, 3 months ]: Measured using a craving questionnaire for cocaine validated in Mexican population (Cocaine Craving Questionnaire or CCQ).
2. Change in Cocaine Craving (VAS) [ Time Frame: Baseline, 2 weeks, 3 months ]: Measured using a 100 mm visual analog scale (VAS).
3. Change in Cocaine Urine Test [ Time Frame: Baseline, 2 weeks, 3 months ]: Frequency of cocaine use measured using reagent strips from Instant View drug screening (> 300 ng/mL). Results are Positive or Negative.

- Secondary outcome measures:

1. Changes in Psychopathological Symptoms [ Time Frame: Baseline, 2 weeks, 3 months ]: Measured by the 90 Symptoms Questionnaire (SCL-90).
2. Changes in Depression [ Time Frame: Baseline, 2 weeks, 3 months ]: Measured by Hamilton Depression Rating Scale (HDRS) (21 items).
3. Changes in Anxiety [ Time Frame: Baseline, 2 weeks, 3 months ]: Measured by Hamilton Anxiety Rating Scale (HARS).

4. Changes in Drug Consumption and Related Problems [ Time Frame: Baseline, 2 weeks, 3 months ]: Measured by the Addiction Severity Index (ASI-lite).
5. Changes in Sleep Quality: PSQI [ Time Frame: Baseline, 2 weeks, 3 months ]: Measured by the Pittsburgh Sleep Quality Index (PSQI).
6. Changes in Impulsivity [ Time Frame: Baseline, 2 weeks, 3 months ]: Measured by the Barratt Impulsivity Scale-11 (BIS-11).
7. Lapse rate [ Time Frame: Baseline, 2 weeks, 3 months ]: Lapse is defined as at least one consumption event not in the same pattern as the baseline consumption. The report of self-consumption of cocaine and urine drug tests, with special attention to the presence of traces of cocaine.
8. Relapse rate [ Time Frame: Baseline, 2 weeks, 3 months ]: Relapse is defined as consumption events in the same pattern as the baseline consumption. The report of self-consumption of cocaine and urine drug tests, with special attention to the presence of traces of cocaine.

- Tertiary outcome measure:

1. Changes in resting state functional connectivity using magnetic resonance imaging

**Clinical statistical analysis**

All baseline tables were constructed using R toolbox moonBook ([https://github.com/cardiomoon/moonBook](https://github.com/cardiomoon/moonBook)), which performs Shapiro-Wilks test for normality, then provides a statistical test for 2 independent groups as follows: 1) continuous normal = unpaired t-test, 2) continuous not normal = Wilcoxon test, 3) nominal = chi-square or Fisher's test, with 2-tailed alpha 0.05. Longitudinal tables were also constructed with moonBook, however statistical tests were independent of the toolbox. Group and time interaction of primary and secondary outcome continuous variables in the acute phase were analyzed with a 2 x 2 Mixed Model in R and RStudio (lme4), with craving (CCQ-Now and VAS) or impulsivity as dependent variables, age and sex as covariates (fixed effects) and patients as a random effect. For the mixed models we calculated effect sizes using Cohen's d and the EMAtools package, with correction for samples smaller than 50 subjects. Urine tests for cocaine (nominal) were analyzed using the Cochran-Mantel-Haenszel test (15,16) and classifying individual trajectories into four patterns: 1) Maintained negative results, 2) maintained positive results, 3) changed from negative to positive, and 4) changed from positive to negative. We expected the Active group would have the highest percentages of the first and fourth patterns. Finally, correlation was followed by a multiple regression analysis using all demographic measures, years of cocaine use, craving (CCQ-Now and VAS) and impulsivity, with the delta correlation (functional connectivity) being the dependent variable. We used a stepwise regression with both forward and backward selection for searching the minimum Akaike Information Criterion (AIC) which provides a means for model selection, providing the loss of information for each model tested after removing the multicollinearity.

For the open maintenance phase, we pooled subjects from both groups regardless of initial assignment (Sham and Active). Primary and secondary outcome measures were analyzed with a Mixed Model using the *lme4* package and age and sex as covariates, with unequal sample sizes between time points, and the *multcomp* package for post hoc contrasts. Urine test for cocaine (nominal) was analyzed using Cochran's Q test (15), so it could only be computed for the 15 subjects with complete urine tests up to 6 months. All longitudinal tests were corrected for multiple comparisons when a similar test was repeated, using false discovery rate (FDR) at q < 0.05, with the number of tests grouped into either primary n = 2 or secondary outcomes n = 4. For *post hoc* contrasts in mixed models we used the Tukey correction. For the mTLFB we used the Friedman test on the sample that completed 6 months (n = 15), and *post hoc* contrasts using the Wilcox test corrected with FDR for the variables *frequency of use per month* and *grams per month*. Effect size was calculated with Kendall's W.

**Transcranial magnetic stimulation**

We performed a double-blind randomized controlled trial (RCT) with parallel groups (Sham/Real) with a final allocation ratio of 1:1.25 for 2 weeks of acute treatment named the *acute phase*, following with an open-label trial at timepoints 3, 6 and up to 12 months of chronic treatment maintenance, named the *maintenance phase*. The allocation was 1:1, however it would have been simple for TMS technicians to guess the group allocation for the last patients as they knew the final sample size and group membership of previous patients. Therefore, we decided to include a bigger sample for the randomization to avoid guessing of the group. For the acute phase, we used a MagPro R30+Option magnetic stimulator and an eight-shaped B65-A/P coil (Magventure, Denmark), and for the maintenance phase, we used a MagPro R30 stimulator and an eight-shaped MCF-B70 (Magventure, Denmark). The reason for using 2 different TMS models was practical, to be able to stimulate more patients. However, there are no differences in the induced field between models, only the cooling system and the sham possibility from the MagPro R30+Option. Also, the eight-shaped coil B65-A/P coil is angled while the eight-shaped MCF-B70 is flat. We used a 5-Hz excitatory frequency as it is standard in our clinical setting due to the low presence of secondary effects and similar clinical improvement to 10-Hz in major depression, Alzheimer's disease, among others (17–21). Safety outcomes are reported in Table S3. The motor threshold was determined in each patient as described by Rossini et al. (22), localizing M1 from vertex 5 cm along and 2 cm anteriorly the interaural line. The coil was placed at 45° with respect to the interhemispheric fissure (anterior-medial induced current) and single pulses were applied separated by 5 seconds. The intensity that caused at least 5 responses of the abductor *pollicis brevis* (APB) muscle from 10 pulses was considered the MT (23). MT was determined before the first session and on the 6th day of treatment. For the maintenance phase, MT was determined in each session (once per week). We localized left DLPFC using the 5 cm method in the first 16 participants and the Beam F3 method (Beam, Borckardt, Reeves, & George, 2009) in the rest of the subjects to optimize DLPFC localization (only n = 11 were available at the time for this analysis). Sham electrodes were placed to simulate muscle contraction in the Sham group. The acute phase comprised 10 weekdays of 5,000 pulses per day (two sessions of 50 trains at 5 Hz, 50 pulses/train, 10 s inter-train interval and 15 min inter-session interval). The maintenance phase comprised 3 and 6 months of 5,000 pulses per day, 2 sessions per week. The motor threshold was maintained at 100% in all patients. Because a Brain Navigator was not available, we used a vitamin E capsule fiducial during MRI acquisition to identify the actual stimulation target where rTMS was delivered in n = 27. EMS oversaw all rTMS sessions and determined the capsule's location before the first MRI session using either the 5.5 cm anatomic criterion or the Beam F3 method (Table S4 & Fig. S2). We changed to the superior Beam F3 method after the first 16 participants to improve lDLPFC localization (24). EMS marked lDLPFC on the scalp with a marker, then maintained the capsule's position using removable tape and a swimmer's cap. Subsequently, EMS checked the capsule location before scanning. That same marked location on the scalp based on the coordinates at which the fucidal (capsule) was placed was used for rTMS sessions.

**Table S3.** Safety outcomes for the acute phase.

| | SHAM (N=240) | ACTIVE (N=300) | p |
|---|---|---|---|
| Headache | | | 0.026 |
| -0 | 216 (97.7%) | 244 (90.7%) | |
| -1 | 1 ( 0.5%) | 5 ( 1.9%) | |
| -2 | 0 ( 0.0%) | 10 ( 3.7%) | |
| -3 | 3 ( 1.4%) | 7 ( 2.6%) | |
| -4 | 1 ( 0.5%) | 2 ( 0.7%) | |
| -5 | 0 ( 0.0%) | 1 ( 0.4%) | |
| Neck pain | | | 0.058 |
| -0 | 188 (85.1%) | 242 (90.0%) | |
| -1 | 1 ( 0.5%) | 5 ( 1.9%) | |
| -2 | 18 ( 8.1%) | 17 ( 6.3%) | |
| -3 | 11 ( 5.0%) | 4 ( 1.5%) | |
| -4 | 3 ( 1.4%) | 1 ( 0.4%) | |
| -5 | 0 (0.0%) | 0 (0.0%) | |
| Scalp pain | | | 0.17 |
| -0 | 219 (99.1%) | 261 (97.0%) | |
| -1 | 2 ( 0.9%) | 2 ( 0.7%) | |
| -2 | 0 ( 0.0%) | 5 ( 1.9%) | |
| -3 | 0 ( 0.0%) | 1 ( 0.4%) | |
| -4 | 0 (0.0%) | 0 (0.0%) | |
| -5 | 0 (0.0%) | 0 (0.0%) | |

| | | | |
|---|---|---|---|
| Cognitive decline | | | 0.567 |
| -0 | 221 (100.0%) | 267 (99.3%) | |
| -1 | 0 (0.0%) | 0 (0.0%) | |
| -2 | 0 ( 0.0%) | 2 ( 0.7%) | |
| -3 | 0 (0.0%) | 0 (0.0%) | |
| -4 | 0 (0.0%) | 0 (0.0%) | |
| -5 | 0 (0.0%) | 0 (0.0%) | |
| Concentration decline | | | 0.346 |
| -0 | 221 (100.0%) | 265 (98.5%) | |
| -1 | 0 ( 0.0%) | 1 ( 0.4%) | |
| -2 | 0 ( 0.0%) | 1 ( 0.4%) | |
| -3 | 0 ( 0.0%) | 2 ( 0.7%) | |
| -4 | 0 (0.0%) | 0 (0.0%) | |
| -5 | 0 (0.0%) | 0 (0.0%) | |
| Hearing decline | | | 0.479 |
| -0 | 221 (100.0%) | 266 (98.9%) | |
| -1 | 0 ( 0.0%) | 1 ( 0.4%) | |
| -2 | 0 ( 0.0%) | 1 ( 0.4%) | |
| -3 | 0 (0.0%) | 0 (0.0%) | |
| -4 | 0 ( 0.0%) | 1 ( 0.4%) | |
| -5 | 0 (0.0%) | 0 (0.0%) | |
| Irritation | | | 0.053 |
| -0 | 218 (98.6%) | 259 (96.3%) | |
| -1 | 0 ( 0.0%) | 7 ( 2.6%) | |

| | | | |
|---|---|---|---|
| -2 | 3 ( 1.4%) | 3 ( 1.1%) | |
| -3 | 0 (0.0%) | 0 (0.0%) | |
| -4 | 0 (0.0%) | 0 (0.0%) | |
| -5 | 0 (0.0%) | 0 (0.0%) | |
| Seizures | | | |
| -0 | 221 (100.0%) | 269 (100.0%) | |
| Mood changes | | | 0.361 |
| -0 | 220 (99.5%) | 269 (100.0%) | |
| -1 | 0 (0.0%) | 0 (0.0%) | |
| -2 | 1 ( 0.5%) | 0 ( 0.0%) | |
| -3 | 0 (0.0%) | 0 (0.0%) | |
| -4 | 0 (0.0%) | 0 (0.0%) | |
| -5 | 0 ( 0.0%) | 0 ( 0.0%) | |

*Scale from 0 = none to 5 = severe. 10 sessions per patient before attrition (Sham n = 24, Active n= 30).*

**Table S4.** Type of lDLPFC localization per patient.

| ID | Group | Method | MNIx | MNIy | MNIz |
|---|---|---|---|---|---|
| 2 | Sham | 5.5 cm | -20 | 62 | 26 |
| 4 | Active | 5.5 cm | -16 | 52 | 40 |
| 8 | Sham | 5.5 cm | -24 | 48 | 42 |
| 20 | Active | 5.5 cm | -28 | 46 | 42 |
| 21 | Active | 5.5 cm | -24 | 50 | 36 |
| 22 | Sham | 5.5 cm | -36 | 50 | 28 |
| 23 | Active | 5.5 cm | -30 | 42 | 40 |
| 24 | Active | 5.5 cm | -34 | 36 | 44 |
| 25 | Active | 5.5 cm | -26 | 42 | 46 |
| 26 | Active | 5.5 cm | -30 | 48 | 36 |
| 27 | Sham | 5.5 cm | -30 | 50 | 32 |
| 30 | Sham | 5.5 cm | -28 | 52 | 32 |
| 31 | Active | 5.5 cm | -26 | 48 | 36 |
| 32 | Active | 5.5 cm | -28 | 46 | 38 |
| 33 | Sham | 5.5 cm | -22 | 44 | 46 |
| 34 | Active | 5.5 cm | -24 | 50 | 34 |
| 36 | Sham | Beam F3 | -26 | 52 | 32 |

| | | | | | |
|---|---|---|---|---|---|
| 37 | Active | Beam F3 | -36 | 28 | 52 |
| 39 | Active | Beam F3 | -38 | 36 | 40 |
| 41 | Active | Beam F3 | -32 | 54 | 26 |
| 42 | Sham | Beam F3 | -36 | 32 | 48 |
| 43 | Active | Beam F3 | -46 | 32 | 38 |
| 45 | Sham | Beam F3 | -40 | 48 | 30 |
| 46 | Sham | Beam F3 | -32 | 46 | 36 |
| 47 | Sham | Beam F3 | -40 | 42 | 36 |
| 48 | Active | Beam F3 | -40 | 42 | 36 |
| 49 | Active | Beam F3 | -42 | 50 | 26 |

The last 3 columns show the localization of the TMS in each patient in MNI coordinates.

**Figure S2.** TMS target locations between methods.

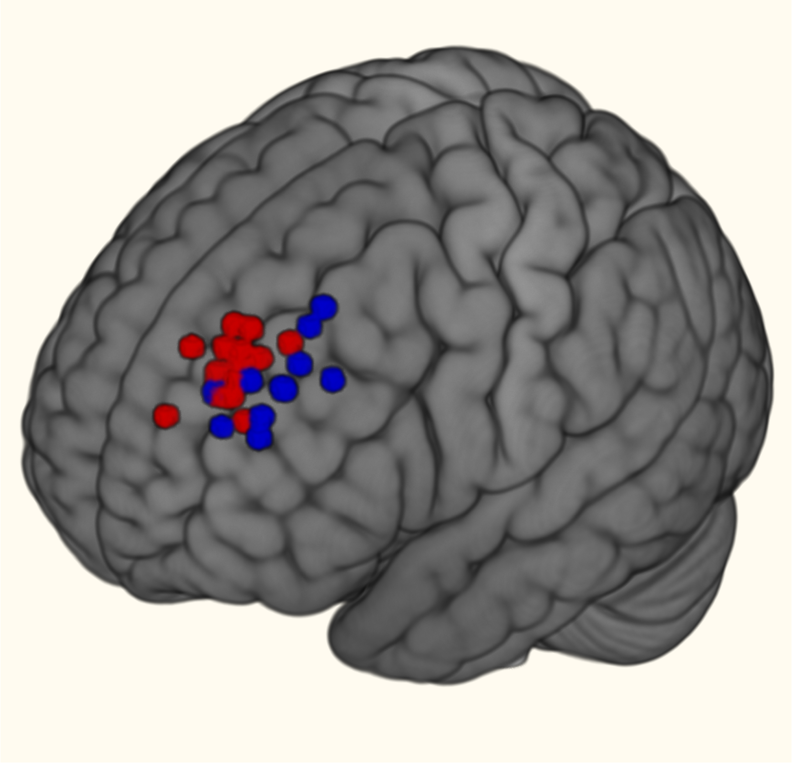

Red = 5.5 cm; blue = Beam F3.

**Sham and double-blind protocol**

Researcher JJGO created the randomization to allocate patients, which was entered into the MagVenture's Research Study System software and saved in a USB memory chip with the protocol selection (Sham or Active rTMS) for each patient and delivered to RAL and EMS for the rTMS sessions. The USB was especially programmed to avoid showing any information to operators to maintain the blinding and only JJGO knew the group membership before the open label. The way the sham works in the stimulator is the following: the programmed USB is inserted and the software asks the TMS technician to place the coil with either side facing down to the scalp, without clues of which side of the coil is stimulating (upward or downward). The technician had no clinical knowledge of the patients to avoid bias. To enhance the sham and blinding, electrodes were placed on each patient on the left temporalis muscle to simulate muscle contraction in the Sham group. Out of 53 patients, the blinding was assessed in 30 of the patients and the results showed 63% of the patients guessed their group allocation correctly, while researchers guessed group allocations 52% correctly.

**Fiducial to standard space**

First, we registered the location of the stimulation region on the scalp, by manually locating the coordinates of the fiducial in the participants' space using fslview. To avoid any distortion in the algorithm, we co-registered a different high-definition structural image to the space of the fiducial image and made a deskulled version of it; this was done with ANTs. Using both the full-head and brain co-registered images, with MATLAB 2019a we located the point most proximal to the cortex in the projection with a 90° angle to the tangent of the head surface in a coronal slice. A single point-seed mask with these cortex coordinates was created for each participant and registered to the standard MNI152 template with ANTs. Finally, we registered the coordinates of all of these normalized stimulation locations with their coordinates in the standard MNI space and with this information calculated the average central stimulation region in the brain cortex.

**TMS regions of interest**

The lDLPFC ROIs were specified as per (25). Briefly, cones with 12 mm radius were centered at each individual stimulation coordinate in MNI (Figure 2, main manuscript). The cones were built with decreasing intensity from the center to the periphery and were based on an approximation of the electric field induced by a standard figure-eight coil. A gray matter mask was used to mask the cones and each cone was normalized to an average value of 1. Normative connectivity was determined using this cone as a weighted mask and n = 1000 subjects from the Human Connectome Project.

**Magnetic resonance imaging**

Neuroimaging data were acquired using a Philips Ingenia 3T scanner (Philips, USA) with a 32-channel Philips head coil. For each MRI session we acquired the following sequences in order: 1) Resting state functional magnetic resonance imaging (rsfMRI), gradient echo planar imaging, TR/TE = 2000/30 ms, FOV = 240 mm, Matrix = 70 x 70, ReconMatrix = 80, slice thickness = 3.33 mm, FA = 75 degrees, voxel = 3 x 3 x 3.33 mm, axial, slices = 37, direction = AP, 2) Structural T1w 3D FFE Sagital, TR/TE = 7/3.5 ms, FA = 8 degrees, FOV = 240 mm, matrix = 240 x 240, voxel = 1 x 1 x 1 mm, gap = 0. To correct for field inhomogeneities we acquired a rsfMRI sequence with 5 volumes in the opposite phase-encoding direction (PA). We also acquired a high angular diffusion-weighted imaging (HARDI) sequence not reported here.

**MRI preprocessing**

Image processing was performed with FMRIPREP (26) version 1.5.5 [1, 2, RRID:SCR_016216], a Nipype (27) [3, 4, RRID:SCR_002502] based tool. Each T1w (T1-weighted) volume was corrected for INU (intensity non-uniformity) using N4BiasFieldCorrection v2.1.0 (28) and skull-stripped using antsBrainExtraction.sh v2.1.0 (using the OASIS template). Spatial normalization to the ICBM 152 Nonlinear Asymmetrical template version 2009c [7, RRID:SCR_008796] was performed through nonlinear registration with the antsRegistration tool of ANTs v2.1.0 [8, RRID:SCR_004757], using brain-extracted versions of both T1w volume and template. Brain tissue segmentation of cerebrospinal fluid (CSF), white-matter (WM) and gray-matter (GM) was performed on the brain-extracted T1w using fast (29,30) (FSL v5.0.9, RRID:SCR_002823).

Functional data was slice-time corrected using 3dTshift from AFNI v16.2.07 [11, RRID:SCR_005927] (31) and motion corrected using mcflirt (FSL v5.0.9 [9]) (32). Distortion correction was performed using an implementation of the TOPUP technique [10] using 3dQwarp (AFNI v16.2.07 [11]). This was followed by co-registration to the corresponding T1w using boundary-based registration [16] with six degrees of freedom, using flirt (FSL) (33). Motion correcting transformations, field distortion correcting warp, BOLD-to-T1w transformation and T1w-to-template (MNI) warp were concatenated and applied in a single step using antsApplyTransforms (ANTs v2.1.0) using Lanczos interpolation.

Physiological noise regressors were extracted applying CompCor (34). Principal components were estimated for the two CompCor variants: temporal (tCompCor) and anatomical (aCompCor). A mask to exclude signals with cortical origin was obtained by eroding the brain mask, ensuring it only contained subcortical structures. Six tCompCor components were then calculated including only the top 5% variable voxels within that subcortical mask. For aCompCor, six components were calculated within the intersection of the subcortical mask and the union of CSF and WM masks calculated in T1w space, after their projection to the native space of each functional run. Framewise displacement was calculated for each functional run using the implementation of Nipype. Many internal operations of FMRIPREP use Nilearn [22, RRID:SCR_001362], principally within the BOLD-processing workflow. For more details of the pipeline see *https://fmriprep.readthedocs.io/en/stable/workflows.html*.

Post-processing was done using XCP Engine, which is a free and open-source software package with a modular design that incorporates the functions of several neuroimaging analysis tools (35,36). It has an optimized confound regression procedure to reduce the influence of subject motion (37,38). The code for the whole pipeline is freely and publicly available in the repo (https://github.com/PennBBL/xcpEngine). The output data from FMRIPREP (i.e. both the minimally preprocessed Images and the functional nuisance regressors matrices) were fed into XCP and preprocessed following the regression strategy of 36 parameters and scrubbing (39), which included: i) correction for distortions induced by inhomogeneities in the magnetic field, ii) removal of the 4 initial volumes, iii) realignment of all volumes to a selected reference volume, iv) demeaning and removal of linear and quadratic trends, v) co-registration of functional data to the high-resolution structural image, vi) removal of nine confounding signals (six motion parameters; global, white matter and cerebrospinal fluid) as well as their derivatives, quadratic terms, and squares of derivatives, vii) motion scrubbing, as in, the removal of every volume that surpassed the motion threshold of 0.5 mm of framewise displacement, as well as both contiguous volumes; and, viii) temporal filtering with a first-order Butterworth filter using a bandpass between 0.01 and 0.08 Hz.

**Functional connectivity analysis**

Because we were interested in the effects of rTMS on lDLPFC-vmPFC FC, we first determined the regions of interest (ROIs) of the lDLPFC circuit by obtaining its normative FC using the methods described in Weigand et al. (25). Briefly, we obtained the MNI coordinates for each subject's stimulated region using its individual fiducial location (n = 27) (Figure 2, main manuscript). We then calculated the average MNI coordinates of lDLPFC stimulation (x = -31,y = 46, z = 36) which was fed to an in-house pipeline by members of the M. D. Fox laboratory to obtain the normative lDLPFC FC map. The pipeline creates a "cone" shaped mask that emulates the induced electric field's decreasing intensity from center to periphery on the cortex when using the standard figure-eight coil. From that weighted mask, correlation maps were calculated in Human Connectome Project fMRI data (n = 1000). The final normative lDLPFC FC map was then clustered using FSL with a Pearson's correlation threshold of r = ± 0.2 and a minimum cluster size of 10 voxels. The cluster corresponding to vmPFC (Figure S3) was used to determine its connectivity with the stimulated lDPFC and potential changes in response to rTMS. Other clusters found included bilateral DLPFC, frontal pole, posterior cingulate cortex (PCC), precuneus, anterior cingulate cortex (ACC), bilateral insula, left striatum, bilateral hippocampus, middle temporal gyrus, supramarginal gyrus and bilateral cerebellum, which were used for exploratory analyses (Table S5).

To calculate lDLPFC FC in each subject and session, we used the average lDLPFC cone mask in FSL on the individual 4D residual volumes, which has yielded similar results to using individualized masks (40). Because our primary interest was lDLPFC - vmPFC FC, the *acute phase* was analyzed with a 2 x 2 Mixed Model (group by session interaction) using FSL randomise at alpha 0.05 with TFCE correction, with the vmPFC cluster as our ROI. Afterwards, we explored the remaining cluster ROIs with the same model. Depending on the results of the clinical and MRI analyses, we correlated the significant FC clusters with significant clinical variables using their delta scores (T1-T0) and Pearson's correlation at alpha 0.05. The maintenance phase was analyzed using the mean FC from the resulting cluster or clusters in the acute phase analysis using a Mixed Model in R (lme4) to account for different sample sizes up to 6 months. Post-hoc, we calculated the individual FC maps of the significant vmPFC cluster (as seed) from the acute phase analysis, and performed a whole-brain 2 x 2 mixed model analysis (group by session) of the acute phase and the maintenance phase.

**Table S4.** Normative Left DLPFC average stimulation cone seed map.

| Hemisphere | Brain region | Voxels | Peak r-value | Peak MNI coordinates | | |
|---|---|---|---|---|---|---|
| | | | | ***x*** | ***y*** | ***z*** |
| **Positive** | | | | | | |
| Left | Dorsolateral prefrontal cortex | 3179 | 0.913 | -30 | 44 | 38 |
| Left | Anterior cingulate cortex | 3170 | 0.465 | -4 | 18 | 36 |
| Left | Anterior insula | 1317 | 0.442 | -34 | 14 | 8 |
| Left | Supramarginal gyrus | 1036 | 0.369 | -62 | -38 | 34 |
| Left | Superior frontal gyrus | 457 | 0.395 | -16 | 6 | 70 |
| Left | Cerebellum VI | 169 | 0.275 | -34 | -50 | -32 |
| Left | Middle frontal gyrus | 67 | 0.286 | -26 | 44 | -12 |
| Left | Precuneus | 32 | 0.217 | -10 | -74 | 42 |
| Left | Cerebellum VIIb | 20 | 0.215 | -40 | -42 | -48 |
| Left | Putamen | 17 | 0.216 | -20 | 14 | -2 |
| Right | Dorsolateral prefrontal cortex | 1858 | 0.547 | 32 | 48 | 32 |
| Right | Midcingulate cortex | 1202 | 0.353 | 12 | -32 | 42 |
| Right | Anterior insula | 1105 | 0.394 | 36 | 16 | 8 |
| Right | Supramarginal gyrus | 805 | 0.352 | 62 | -34 | 36 |
| Right | Cerebellum VI | 194 | 0.279 | 36 | -48 | -32 |
| Right | Cerebellum VIIIa | 80 | 0.235 | 38 | -44 | -52 |
| **Negative** | | | | | | |
| Left | Superior lateral occipital cortex/Angular gyrus | 378 | -0.238 | -50 | -66 | 30 |
| Left | Anterior middle temporal gyrus | 235 | -0.245 | -60 | -8 | -14 |
| Left | Hippocampus | 36 | -0.227 | -24 | -16 | -18 |
| Right | Ventromedial prefrontal cortex | 925 | -0.307 | 2 | 52 | -12 |
| Right | Posterior cingulate cortex/Precuneous | 902 | -0.273 | 2 | -56 | 26 |
| Right | Anterior middle temporal gyrus | 322 | -0.264 | 62 | -4 | -20 |
| Right | Superior lateral occipital cortex/Angular gyrus | 300 | -0.254 | 52 | -60 | 32 |

| | | | | | | |
|---|---|---|---|---|---|---|
| Right | Hippocampus | 58 | -0.238 | 26 | -14 | -18 |
| Right | Middle temporal gyrus | 45 | -0.22 | 66 | -34 | -8 |
| Right | Inferior frontal gyrus | 35 | -0.231 | 40 | 38 | -14 |

*R = Pearson's r.*

**Figure S3.** vmPFC region-of-interest mask.

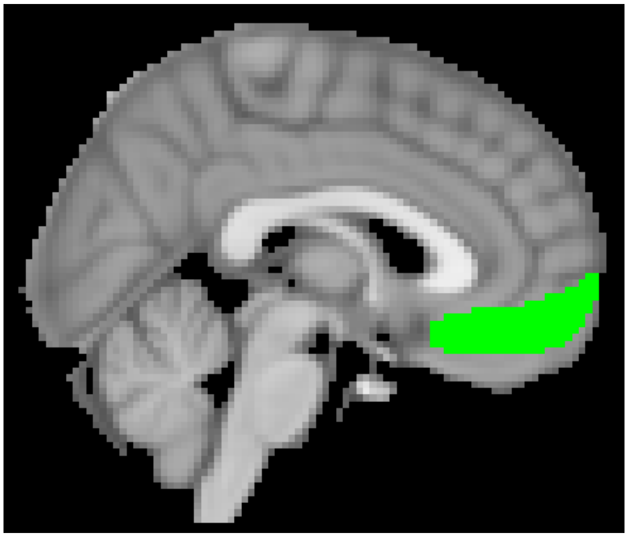

*Sagittal view in FSL MNI152 T1w 2mm template.*

### Supplementary Results

**Table S6.** Urine test results of the acute phase.

| | T0 | | | T1 | | |
|---|---|---|---|---|---|---|
| | **SHAM** (N=20) | **ACTIVE** (N=24) | p-value | **SHAM** (N=20) | **ACTIVE** (N=24) | p-value |
| Cocaine | | | 0.89 | | | 1 |
| - positive | 7 (35.0%) | 10 (41.7%) | | 9 (45.0%) | 10 (41.7%) | |
| - negative | 13 (65.0%) | 14 (58.3%) | | 11 (55.0%) | 14 (58.3%) | |
| Amphetamines | | | | | | |
| - positive | 0 ( 0.0%) | 0 ( 0.0%) | | 0 ( 0.0%) | 0 ( 0.0%) | |
| - negative | 20 (100.0%) | 22 (100.0%) | | 19 (100.0%) | 24 (100.0%) | |
| Benzodiazepines | | | 0.75 | | | 1 |
| - positive | 2 (10.0%) | 4 (18.2%) | | 3 (15.8%) | 3 (12.5%) | |
| - negative | 18 (90.0%) | 18 (81.8%) | | 16 (84.2%) | 21 (87.5%) | |
| Methamphetamines | | | | | | |
| - positive | 0 ( 0.0%) | 0 ( 0.0%) | | 0 ( 0.0%) | 0 ( 0.0%) | |
| - negative | 20 (100.0%) | 22 (100.0%) | | 19 (100.0%) | 24 (100.0%) | |
| Opioids | | | | | | |
| - positive | 0 ( 0.0%) | 0 ( 0.0%) | | 0 ( 0.0%) | 0 ( 0.0%) | |
| - negative | 20 (100.0%) | 22 (100.0%) | | 19 (100.0%) | 24 (100.0%) | |
| Cannabis | | | 1 | | | 0.79 |
| - positive | 5 (25.0%) | 5 (20.8%) | | 4 (20.0%) | 3 (12.5%) | |

| | | | | |
|---|---|---|---|---|
| - negative | 15 (75.0%) | 19 (79.2%) | 16 (80.0%) | 21 (87.5%) |

**Table S7.** Urine test results of the maintenance phase.

| | **BASELINE** (N=41) | **2W** (N=37) | **3M** (N=19) | **6M** (N=14) |
|---|---|---|---|---|
| Cocaine | | | | |
| - positive | 17 (41.5%) | 16 (43.2%) | 6 (31.6%) | 4 (28.6%) |
| - negative | 24 (58.5%) | 21 (56.8%) | 13 (68.4%) | 10 (71.4%) |
| Amphetamines | | | | |
| - positive | 0 ( 0.0%) | 0 ( 0.0%) | 0 ( 0.0%) | 1 ( 7.1%) |
| - negative | 41 (100.0%) | 37 (100.0%) | 19 (100.0%) | 13 (92.9%) |
| Benzodiazepines | | | | |
| - positive | 7 (17.1%) | 4 (10.8%) | 5 (26.3%) | 3 (21.4%) |
| - negative | 34 (82.9%) | 33 (89.2%) | 14 (73.7%) | 11 (78.6%) |
| Methamphetamines | | | | |
| - positive | 0 ( 0.0%) | 0 ( 0.0%) | 0 ( 0.0%) | 1 ( 7.1%) |
| - negative | 41 (100.0%) | 37 (100.0%) | 19 (100.0%) | 13 (92.9%) |
| Opioids | | | | |
| - positive | 0 ( 0.0%) | 0 ( 0.0%) | 0 ( 0.0%) | 0 ( 0%) |
| - negative | 41 (100.0%) | 37 (100.0%) | 19 (100.0%) | 14 (100.0%) |

| | | | | |
|---|---|---|---|---|
| Cannabis | | | | |
| - positive | 8 (19.5%) | 5 (13.5%) | 0 ( 0.0%) | 0 ( 0.0%) |
| - negative | 33 (80.5%) | 32 (86.5%) | 19 (100.0%) | 14 (100.0%) |

**Table S8.** Cocaine urine tests in the acute phase.

| | Group | | | | |
|---|---|---|---|---|---|
| | Sham | | | Active | |
| **ID** | T0 | T1 | **ID** | T0 | T1 |
| *1* | green | green | *4* | green | green |
| *2* | green | green | *7* | green | green |
| *6* | green | green | *15* | green | green |
| *8* | green | green | *20* | green | green |
| *10* | green | green | *23* | green | green |
| *12* | green | green | *31* | green | green |
| *19* | green | green | *34* | green | green |
| *22* | green | green | *39* | green | green |
| *27* | green | green | *41* | green | green |
| *45* | green | green | *43* | green | green |
| *47* | green | green | *48* | green | green |
| *50* | green | green | *54* | green | green |
| *13* | red | red | *3* | red | red |
| *30* | red | red | *11* | red | red |
| *42* | red | red | *16* | red | red |
| *46* | red | red | *24* | red | red |
| *52* | red | red | *37* | red | red |
| *33* | red | green | *49* | red | red |
| *18* | green | red | *17* | red | green |
| *36* | green | red | *25* | red | green |
| | | | *32* | red | green |
| | | | *21* | green | red |
| | | | *26* | green | red |
| | | | *53* | green | red |

*T0 = Baseline, T1 = 2 weeks of rTMS treatment, ID = identificatioon, red = positive for cocaine, green = negative for cocaine.*

**Table S9.** Cocaine urine tests in the maintenance phase.

| ID | Time point: Baseline | 2 Weeks | 3 Months | 6 Months |
|---|---|---|---|---|
| 4 | green | green | green | green |
| 7 | green | green | green | green |
| 8 | green | green | green | green |
| 15 | green | green | NA | NA |
| 20 | green | green | green | green |
| 22 | green | green | green | green |
| 23 | green | green | green | green |
| 27 | green | green | green | green |
| 41 | green | green | green | green |
| 3 | red | red | red | red |
| 24 | red | red | red | red |
| 36 | red | red | red | red |
| 25 | red | green | green | green |
| 32 | red | green | green | green |
| 2 | green | green | green | red |

*ID = Identification, red = positive for cocaine, green = negative for cocaine.*

**Table S10.** Maintenance phase multiple comparisons with Tukey correction.

| Contrast | Estimate | Std. Error | z-value | p-value |
|---|---|---|---|---|
| CCQ-Now | | | | |
| 2W - BASELINE | -23.24 | 6.94 | -3.35 | 0.00 |
| 3M - BASELINE | -35.48 | 8.77 | -4.05 | 0.00 |
| 6M - BASELINE | -28.92 | 9.81 | -2.95 | 0.02 |
| 3M - 2W | -12.24 | 8.85 | -1.38 | 0.50 |
| 6M - 2W | -5.68 | 9.88 | -0.57 | 0.94 |
| 6M - 3M | 6.56 | 10.65 | 0.62 | 0.93 |
| VAS | | | | |
| 2W - BASELINE | -2.01 | 0.48 | -4.20 | 0.00 |
| 3M - BASELINE | -2.96 | 0.60 | -4.91 | 0.00 |
| 6M - BASELINE | -0.92 | 0.67 | -1.36 | 0.52 |
| 3M - 2W | -0.95 | 0.61 | -1.56 | 0.40 |
| 6M - 2W | 1.09 | 0.68 | 1.61 | 0.37 |
| 6M - 3M | 2.04 | 0.74 | 2.78 | 0.03 |
| Impulsivity | | | | |
| 2W - BASELINE | -9.51 | 2.68 | -3.55 | 0.00 |
| 3M - BASELINE | -13.40 | 3.40 | -3.94 | 0.00 |

| | | | | |
|---|---|---|---|---|
| 6M - BASELINE | -8.45 | 3.80 | -2.22 | 0.11 |
| 3M - 2W | -3.89 | 3.42 | -1.14 | 0.66 |
| 6M - 2W | 1.07 | 3.82 | 0.28 | 0.99 |
| 6M - 3M | 4.95 | 4.11 | 1.21 | 0.62 |
| Anxiety | | | | |
| 2W - BASELINE | -6.12 | 1.82 | -3.36 | 0.00 |
| 3M - BASELINE | -6.01 | 2.30 | -2.61 | 0.04 |
| 6M - BASELINE | -3.06 | 2.57 | -1.19 | 0.63 |
| 3M - 2W | 0.11 | 2.32 | 0.05 | 1.00 |
| 6M - 2W | 3.06 | 2.59 | 1.18 | 0.63 |
| 6M - 3M | 2.95 | 2.80 | 1.05 | 0.71 |
| Depression | | | | |
| 2W - BASELINE | -5.42 | 1.44 | -3.75 | 0.00 |
| 3M - BASELINE | -5.64 | 1.80 | -3.14 | 0.01 |
| 6M - BASELINE | -5.02 | 2.01 | -2.50 | 0.06 |
| 3M - 2W | -0.22 | 1.83 | -0.12 | 1.00 |
| 6M - 2W | 0.40 | 2.03 | 0.20 | 1.00 |
| 6M - 3M | 0.62 | 2.22 | 0.28 | 0.99 |

| Sleep | | | | |
|---|---|---|---|---|
| 2W - BASELINE | -0.98 | 0.76 | -1.29 | 0.56 |
| 3M - BASELINE | -1.08 | 0.93 | -1.15 | 0.65 |
| 6M - BASELINE | -1.83 | 1.04 | -1.76 | 0.29 |
| 3M - 2W | -0.10 | 0.95 | -0.11 | 1.00 |
| 6M - 2W | -0.85 | 1.06 | -0.81 | 0.85 |
| 6M - 3M | -0.75 | 1.15 | -0.65 | 0.91 |

*2W = 2 weeks, 3M = 3 months, 6M = 6 months.*

**Figure S4.** WHODAS and SCL90-R measures in the acute phase.

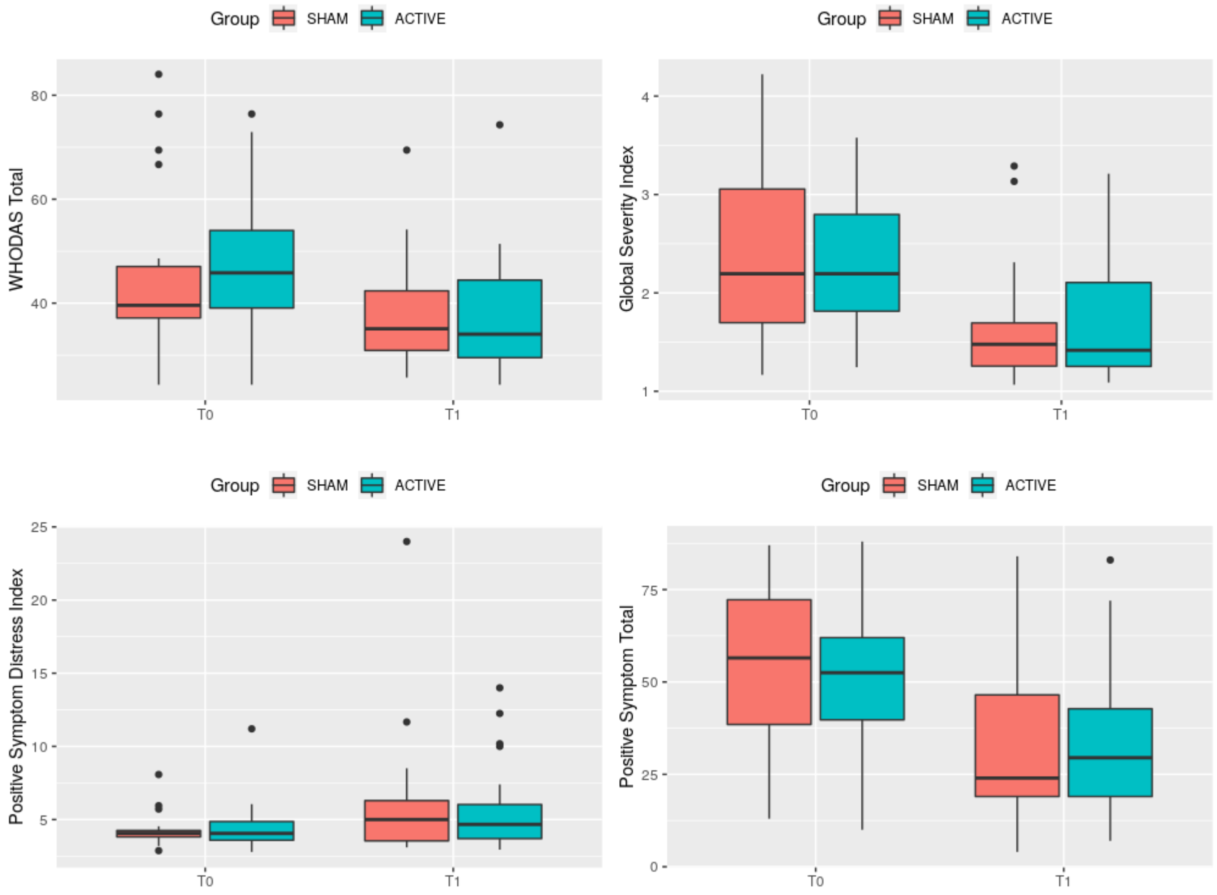

**Table S11.** Other clinical measures in the acute phase.

| | SHAM | | ACTIVE | |
|---|---|---|---|---|
| | T0 (N=20) | T1 (N=20) | T0 (N=24) | T1 (N=24) |
| WHODAS Total | 45.2 ± 16.5 | 38.2 ± 10.9 | 46.8 ± 13.5 | 37.9 ± 11.0 |
| SCL90R - Global Severity Index | 2.3 ± 0.9 | 1.7 ± 0.6 | 2.3 ± 0.6 | 1.7 ± 0.6 |
| SCL90R - Positive Symptom Distress Index | 4.3 ± 1.2 | 6.2 ± 4.8 | 4.5 ± 1.7 | 5.7 ± 3.0 |
| SCL90R - Positive Symptom Total | 53.2 ± 22.2 | 34.4 ± 23.4 | 50.3 ± 19.8 | 34.8 ± 22.6 |

**Figure S5.** WHODAS and SCL90-R measures in the maintenance phase.

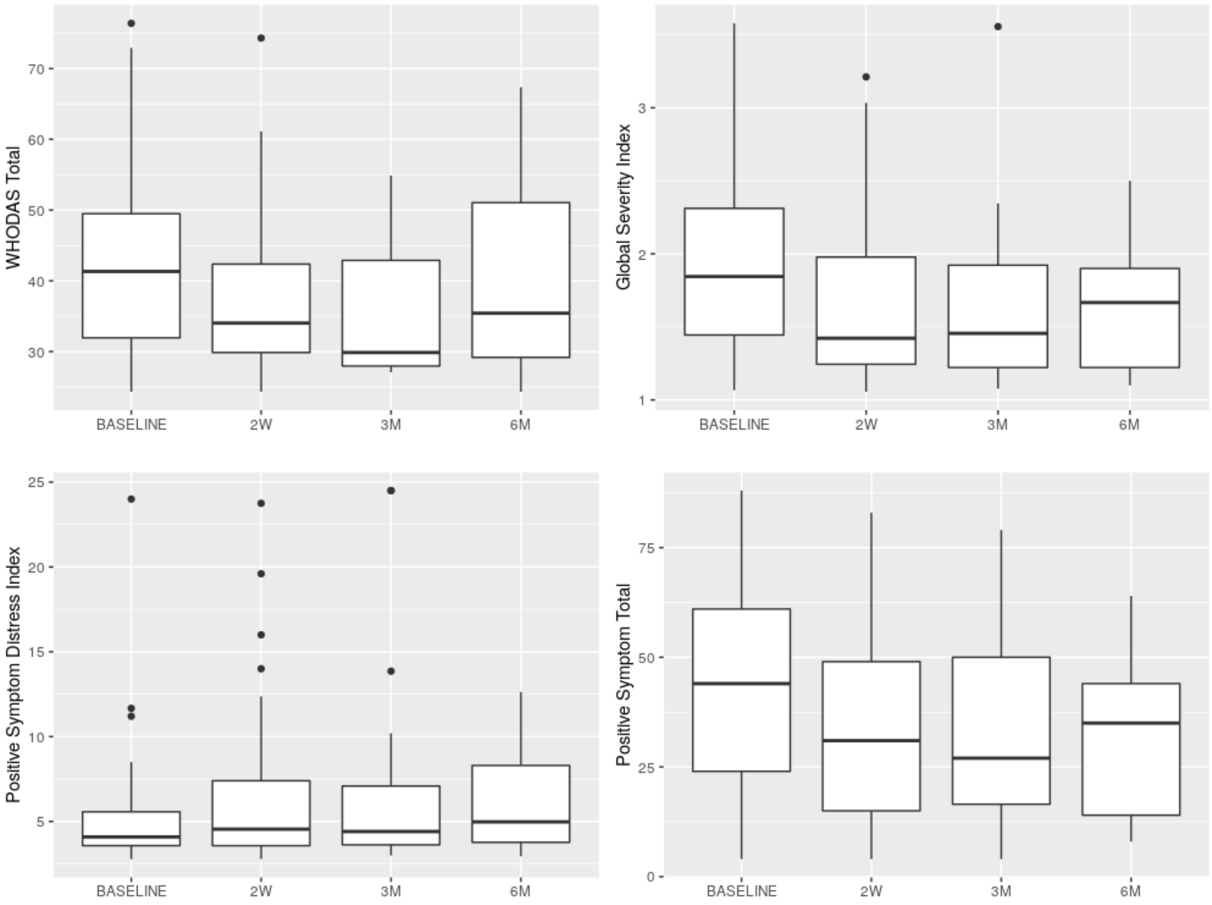

**Table S12.** Other clinical measures in the maintenance phase.

| | BASELINE (N=44) | 2W (N=37) | 3M (N=20) | 6M (N=15) | p |
|---|---|---|---|---|---|
| WHODAS Total | 42.9 ± 13.0 | 36.9 ± 10.3 | 35.9 ± 9.6 | 40.9 ± 14.6 | 0.08 |
| SCL90R - Global Severity Index | 2.0 ± 0.7 | 1.7 ± 0.6 | 1.6 ± 0.6 | 1.6 ± 0.4 | 0.03 |
| SCL90R - Positive Symptom Distress Index | 5.2 ± 3.5 | 6.8 ± 5.0 | 7.6 ± 6.6 | 6.1 ± 3.0 | 0.25 |
| SCL90R - Positive Symptom Total | 43.3 ± 22.7 | 34.0 ± 23.3 | 32.0 ± 22.4 | 31.1 ± 18.9 | 0.12 |

**Table S13.** mTLFB Frequency of use pairwise contrasts.

| Month | Month | n1 | n2 | statistic | p | p.adj | p.adj.signif |
|---|---|---|---|---|---|---|---|
| MONTH-PRE | MONTH-1 | 15 | 15 | 91 | 0.002 | 0.013 | * |
| MONTH-PRE | MONTH-2 | 15 | 15 | 103.5 | 0.002 | 0.013 | * |
| MONTH-PRE | MONTH-3 | 15 | 15 | 100 | 0.003 | 0.013 | * |
| MONTH-PRE | MONTH-4 | 15 | 15 | 100 | 0.003 | 0.013 | * |
| MONTH-PRE | MONTH-5 | 15 | 15 | 114 | 0.002 | 0.013 | * |
| MONTH-PRE | MONTH-6 | 15 | 15 | 98 | 0.005 | 0.016 | * |

| | | | | | | | |
|---|---|---|---|---|---|---|---|
| MONTH-1 | MONTH-2 | 15 | 15 | 13.5 | 0.136 | 0.408 | ns |
| MONTH-1 | MONTH-3 | 15 | 15 | 9 | 0.787 | 0.932 | ns |
| MONTH-1 | MONTH-4 | 15 | 15 | 6 | 0.855 | 0.932 | ns |
| MONTH-1 | MONTH-5 | 15 | 15 | 19.5 | 0.888 | 0.932 | ns |
| MONTH-1 | MONTH-6 | 15 | 15 | 15.5 | 0.866 | 0.932 | ns |
| MONTH-2 | MONTH-3 | 15 | 15 | 2 | 0.789 | 0.932 | ns |
| MONTH-2 | MONTH-4 | 15 | 15 | 2 | 0.789 | 0.932 | ns |
| MONTH-2 | MONTH-5 | 15 | 15 | 8.5 | 0.752 | 0.932 | ns |
| MONTH-2 | MONTH-6 | 15 | 15 | 9 | 0.833 | 0.932 | ns |
| MONTH-3 | MONTH-4 | 15 | 15 | 1.5 | 1 | 1 | ns |
| MONTH-3 | MONTH-5 | 15 | 15 | 9 | 0.786 | 0.932 | ns |
| MONTH-3 | MONTH-6 | 15 | 15 | 4 | 0.85 | 0.932 | ns |
| MONTH-4 | MONTH-5 | 15 | 15 | 12.5 | 0.75 | 0.932 | ns |
| MONTH-4 | MONTH-6 | 15 | 15 | 4 | 0.855 | 0.932 | ns |
| MONTH-5 | MONTH-6 | 15 | 15 | 8 | 0.673 | 0.932 | ns |

mTLFB = Timeline Followback Method Assessment modified; p = p-value; p.adj = FDR adjusted p-value; * = significant.

**Table S14.** mTLFB Grams of cocaine used pairwise contrasts.

| Month | Month | n1 | n2 | statistic | p | p.adj | p.adj.signif |
|---|---|---|---|---|---|---|---|
| MONTH-PRE | MONTH-1 | 15 | 15 | 105 | 0.001 | 0.014 | * |
| MONTH-PRE | MONTH-2 | 15 | 15 | 117 | 0.001 | 0.014 | * |
| MONTH-PRE | MONTH-3 | 15 | 15 | 104 | 0.014 | 0.057 | ns |
| MONTH-PRE | MONTH-4 | 15 | 15 | 101 | 0.021 | 0.075 | ns |
| MONTH-PRE | MONTH-5 | 15 | 15 | 109 | 0.006 | 0.033 | * |
| MONTH-PRE | MONTH-6 | 15 | 15 | 96.5 | 0.006 | 0.033 | * |
| MONTH-1 | MONTH-2 | 15 | 15 | 16 | 0.295 | 0.774 | ns |
| MONTH-1 | MONTH-3 | 15 | 15 | 10 | 1 | 1 | ns |
| MONTH-1 | MONTH-4 | 15 | 15 | 9 | 0.834 | 1 | ns |
| MONTH-1 | MONTH-5 | 15 | 15 | 21 | 0.726 | 1 | ns |
| MONTH-1 | MONTH-6 | 15 | 15 | 14.5 | 1 | 1 | ns |
| MONTH-2 | MONTH-3 | 15 | 15 | 1 | 1 | 1 | ns |
| MONTH-2 | MONTH-4 | 15 | 15 | 1 | 0.423 | 0.987 | ns |
| MONTH-2 | MONTH-5 | 15 | 15 | 7 | 1 | 1 | ns |
| MONTH-2 | MONTH-6 | 15 | 15 | 5 | 0.59 | 1 | ns |
| MONTH-3 | MONTH-4 | 15 | 15 | 0 | 1 | 1 | ns |
| MONTH-3 | MONTH-5 | 15 | 15 | 7 | 0.584 | 1 | ns |

| | | | | | | | |
|---|---|---|---|---|---|---|---|
| MONTH-3 | MONTH-6 | 15 | 15 | 6 | 0.855 | 1 | ns |
| MONTH-4 | MONTH-5 | 15 | 15 | 12 | 0.281 | 0.774 | ns |
| MONTH-4 | MONTH-6 | 15 | 15 | 7 | 0.584 | 1 | ns |
| MONTH-5 | MONTH-6 | 15 | 15 | 8 | 1 | 1 | ns |

mTLFB = Timeline Followback Method Assessment modified; p = p-value; p.adj = FDR adjusted p-value; * = significant.

**Extended study limitations**

Study limitations include substantial dropout, which is common in the treatment of SUDs. A 12-month treatment study found an overall dropout rate of 31%, varying between 15 and 56% across subgroups (41); the same group found 30% dropout in CUD psychological treatment in another study (42). A 69% dropout rate for a 4-week treatment has also been reported in CUD (43). Reasons suggested for dropout included age, education, SUD severity, comorbidities, family problems and work incompatibility (44). Attrition in our study was: 1) 17% at 2 weeks; 2) 63% at 3 months; and 3) 72% at 6 months. The most common reasons were "poor adherence to treatment" followed by "incompatibility due to work." Overall, rTMS as an add-on treatment did not improve adherence beyond standard treatment. CUD patients' motivation to start and adhere to treatment may differ between subjects and timepoints. For example, initial motivation may be to please one's family. However, the urge to consume cocaine often eventually overcomes this initial motivation even in the face of improvement. Motivation to remain in treatment should be measured in future studies to improve dropout rates. Moreover, all our CUD patients had to follow standard treatment, consisting of group psychotherapy, individual psychotherapy and pharmacotherapy. Several of our patients received pharmacotherapy concurrently with starting rMTS treatment but we did not find any effects of pharmacotherapies between active and sham groups. Our double-blind acute phase arm was limited to 2 weeks for ethical reasons, so as not to extend placebo treatment more than necessary for a vulnerable population. Nevertheless, that time window seemed sufficient to detect positive clinical effects. Although 5-Hz of excitatory rTMS was enough to produce significant effects in craving VAS and impulsivity, a stronger effect may be obtainable with 10-20 Hz, as 5-Hz is in the low-range of excitatory frequencies. The lack of neuronavigation is a limitation. However, most outpatient clinics use the 5.5 cm method or the Beam F3 system due to the high cost of neuronavigation. To account for the lack of neuronavigation, we used fucidals in 27 patients to allow for some individual cortical localization in the analysis. Finally, we measured common subjective variables such as craving and impulsivity, and future studies may benefit from using more objective measures such as cue-reactivity (45). The overall goal of our study was to assess rTMS as an add-on therapy to the standard combination of CUD treatment as this is what clinicians will encounter in their daily work. Despite these limitations, we were able to find a positive add-on effect of rTMS in the first 2 weeks.

controlled trial of daily left prefrontal cortex TMS for treating depression. *Biol Psychiatry* 48: 962–970.

32. Jenkinson M, Bannister P, Brady M, Smith S (2002): Improved optimization for the

robust and accurate linear registration and motion correction of brain images. *Neuroimage* 17: 825–841.

40. Cash RFH, Zalesky A, Thomson RH, Tian Y, Cocchi L, Fitzgerald PB (2019):

Subgenual Functional Connectivity Predicts Antidepressant Treatment Response to Transcranial Magnetic Stimulation: Independent Validation and Evaluation of Personalization. *Biol Psychiatry* 86: e5–e7.
